# Predicting the Absence and Gaps in Recorded Blood Pressure Follow-up Among Patients With Hypertension

**DOI:** 10.64898/2026.10.26.26366569

**Authors:** Ronveer Sandhu, Shehr Asghar, Malak Abu Hashish, Jiancheng Ye

## Abstract

**Background:** Electronic health record (EHR) data can help identify patients who would benefit from blood pressure reassessment after an elevated reading.

**Objective:** To develop and internally validate a model to predict the absence of a recorded blood pressure measurement after an elevated reading among adults with hypertension; evaluate the potential yield of targeted outreach and model performance across subgroups; and determine whether recorded contact patterns provide additional information for predicting subsequent cardiovascular hospitalization.

**Methods:** This retrospective cohort study used MIMIC-IV data. Adults with hypertension, a qualifying systolic blood pressure reading >140 mmHg, defined as the first such reading on or after index, and 90 days of subsequent database coverage and survival were included. The primary outcome was the absence of a recorded blood pressure measurement within 90 days of the index reading. Gradient-boosted trees incorporating 62 demographic, clinical, and contact-history features were compared with logistic regression models using a random 6:2:2 training, validation, and test split, with isotonic recalibration. Nested cross-validation was used to assess the tree model. Model discrimination, calibration, simulated outreach yield, and subgroup performance were evaluated.

**Results:** Among 30,942 patients, 10,899 (35.2%) had no recorded blood pressure measurement within 90 days. In the test set (n = 6,189), the gradient-boosted tree model achieved an area under the receiver operating characteristic curve (AUROC) of 0.740 (95% CI, 0.727–0.753), compared with 0.698 for penalized logistic regression and 0.687 for two-variable logistic regression. The calibration slope was 0.91, and the area under the precision–recall curve was 0.568 (95% CI, 0.546–0.593). Selecting the 20% of patients with the highest predicted risk identified 34.2% (95% CI, 32.7–35.7) of patients without a recorded measurement, with a positive predictive value of 60.4% (95% CI, 57.4–63.2). In out-of-fold subgroup analyses using a common selection threshold corresponding to an overall selection rate of 20%, sensitivity was 21.5% among Black patients, 23.3% among Hispanic patients, and 37.2% among White patients. Among 15,200 participants in the landmark analysis, 664 cardiovascular hospitalizations occurred. Increasing and sustained contact patterns were associated with modestly higher hazards of cardiovascular hospitalization, with the association primarily attributable to heart failure hospitalization. For sustained versus minimal contact, the hazard ratio for the composite outcome was 1.40 (95% CI, 1.06–1.85). However, adding contact gaps and trajectories improved the C-statistic by only 0.0023 (95% CI, −0.0006 to 0.0056).

**Conclusions:** The model showed potential for prioritizing blood pressure reassessment outreach, although sensitivity varied across racial and ethnic groups. Recorded contact patterns provided little incremental discrimination for cardiovascular hospitalization.

## INTRODUCTION

Hypertension remains a leading modifiable risk factor for myocardial infarction, stroke, and heart failure [1, 2].Yet blood pressure control remains suboptimal among patients diagnosed with hypertension [3]. Effective management requires repeated engagement with the health system: blood pressure must be measured, antihypertensive medications initiated or adjusted, and elevated readings reassessed.[4] Clinical guidelines recommend reassessment approximately 1 month after treatment initiation or adjustment in response to elevated blood pressure and follow-up every 3 to 6 months once blood pressure is controlled [5]. When elevated readings are not followed by timely reassessment and appropriate management, treatment intensification may be delayed, potentially increasing cardiovascular risk [6].

Reassessment after an elevated reading is therefore a clinically actionable step in hypertension management. In electronic health record (EHR) data, the absence of a subsequent recorded blood pressure measurement may indicate a missed reassessment, although it may also reflect measurement outside the health system. The period following an elevated reading thus represents a potential opportunity for targeted outreach, such as a care-coordinator call to arrange reassessment or follow-up by a nurse or pharmacist. Because clinic outreach capacity is limited, a predictive model could help prioritize patients unlikely to have a recorded reassessment, using information already available in the EHR, including visit history, blood pressure measurements, and prescribing data. Evaluating such models requires more than assessing discrimination. Decision curve analysis can help determine whether model-guided decisions offer net benefit across clinically relevant thresholds [7], while operational evaluation should quantify how many patients without a recorded reassessment could be identified for a given outreach capacity. Equity is also essential because outreach prioritization allocates a limited health care resource.[8] Performance should therefore be assessed across racial and ethnic, insurance, sex, and age groups. Algorithms trained on patterns of health care utilization may reproduce disparities in access rather than differences in clinical need [9]. Moreover, when outcome prevalence differs across groups, a model can be well calibrated within each group yet exhibit unequal sensitivity [10].

Recorded contact patterns may also contain information about subsequent cardiovascular risk, although their interpretation is not straightforward. The concept of retention in care, developed in HIV medicine [11], suggests that patients who disengage from health care may experience worse outcomes. In hypertension care, retention has also been operationalized as a return visit within a specified interval; the Hypertension Treatment in Nigeria Program evaluated follow-up within 37 days of registration [12]. In EHR studies, longitudinal contact has been characterized using trajectory classes [13] or intervals between encounters.[14] However, evidence linking these patterns to cardiovascular outcomes remains limited. Greater continuity of care has been associated with lower cardiovascular risk in national claims data [15], but continuity is conceptually distinct from the frequency of recorded encounters. Studies of EHR-based contact patterns also face several methodological challenges. First, patients receiving care outside the observed health system may be indistinguishable from those who discontinue care; therefore, recorded contact measures utilization within the data source rather than retention in care itself. Second, defining cohorts according to whether patients return for subsequent care conditions inclusion on the behavior being studied and may introduce selection bias [16, 17]. Third, measuring exposure during the same period in which outcomes accrue can introduce immortal time bias [18, 19], while care prompted by an emerging clinical condition may itself generate additional encounters. Finally, because greater illness burden often leads to more frequent health care encounters, higher recorded contact may reflect greater underlying morbidity rather than better care [20].

In this study, we used MIMIC-IV, a deidentified EHR database [21], to develop and internally validate a model predicting the absence of a recorded blood pressure measurement within 90 days after an elevated reading among adults with hypertension. We evaluated its potential to prioritize outreach under fixed clinic-capacity constraints and assessed performance across racial and ethnic, insurance, sex, and age groups. As a secondary objective, we examined whether recorded contact patterns, characterized by trajectory classes and gaps between encounters, were associated with subsequent cardiovascular hospitalization. To establish temporal ordering and reduce bias arising from outcome-related contact and exposure ascertainment, we used a landmark design in which cohort eligibility was defined at the index date, contact patterns were measured before the landmark, and covariates were measured before the index date.

## METHODS

### Data source and study population

We used the hospital module of MIMIC-IV v2.2 [21], which contains hospital admissions, diagnoses, laboratory results, and prescription orders from a single academic medical center in Boston, as well as outpatient measurements, including blood pressure and body mass index. We also used the emergency department module (MIMIC-IV-ED) [22], to identify emergency department visits that did not result in hospital admission and to determine the dates of emergency department contacts. We included adults with an ICD-9 or ICD-10 diagnosis of hypertension and a subsequent outpatient blood pressure measurement (codes are provided in **Table S1**). The index date was the date of the first blood pressure measurement on or after the first hypertension-coded admission. After excluding patients with a surgically correctable secondary cause of hypertension or death within 90 days after index, 48,628 patients were eligible for the study (**Figure 1A**). Cohort entry was based on database coverage rather than subsequent return visits. Some analysis-specific eligibility criteria, however, depended on information observed after the relevant index date: the prediction cohort required database coverage and survival for 90 days after the prediction date, whereas the primary landmark cohort required a record after day 730 to confirm survival (Supplementary Methods; sample denominators for all analyses are reported in **Table S9**). We distinguished three sources of information: database coverage determined cohort entry, evidence of survival determined eligibility for the landmark analysis, and death ascertainment determined the end of follow-up.[23] Death ascertainment in MIMIC-IV extends approximately 1 year beyond the last hospital discharge [21].

**Figure 1.**
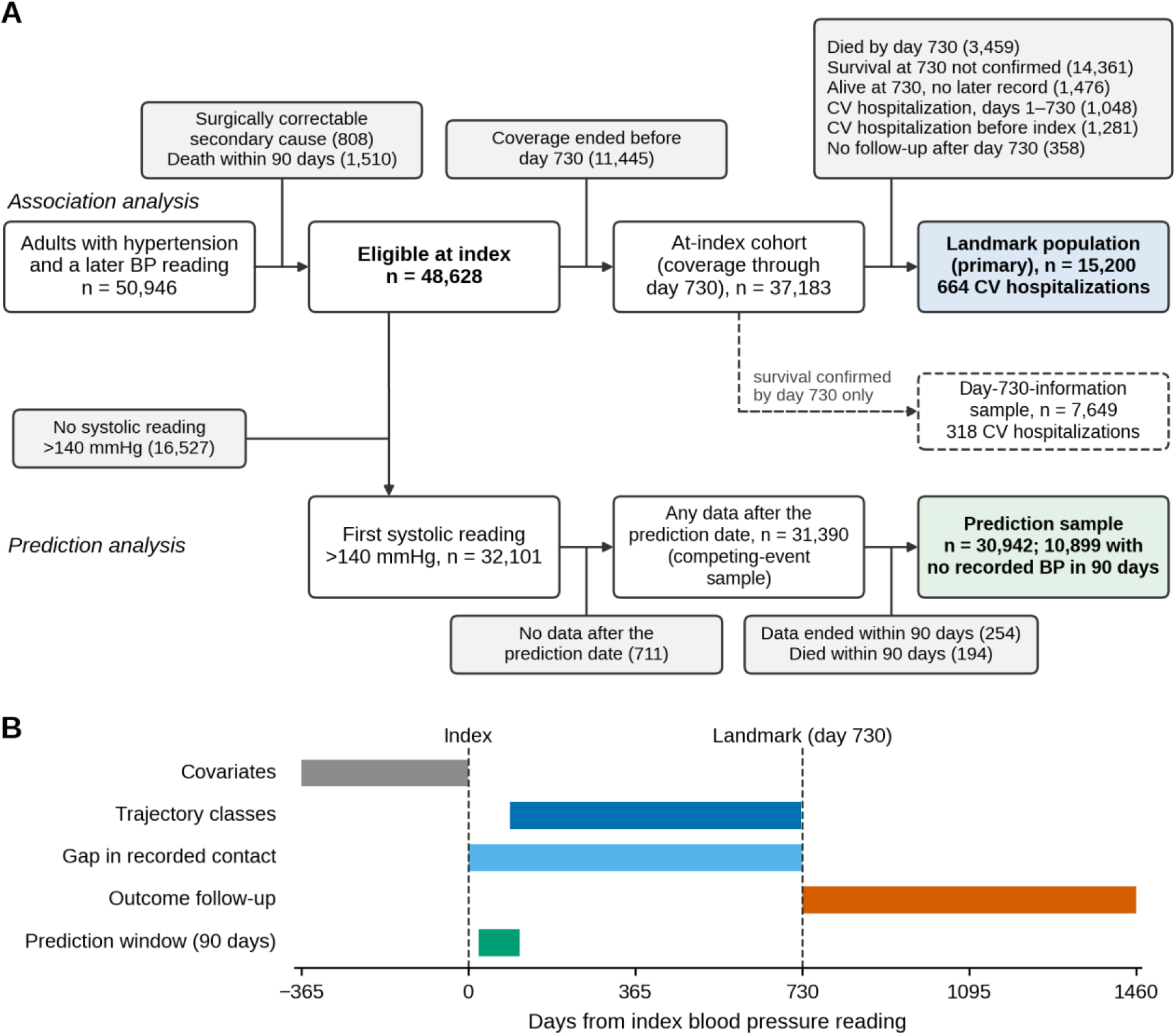
Study population and design. (A) Selection of the association-analysis cohort (top) and prediction-analysis cohort (bottom). Survival at day 730 not confirmed indicates no hospital or emergency department contact during days 366–730 and no record after day 730. Alive at day 730, no later record indicates that survival to the landmark was established but no record after day 730 was available, precluding inclusion in the primary landmark population. The alternative day-730-information sample (dashed outline) confirms survival using only hospital or emergency department contact during days 366–730; 6,432 of its 7,649 patients also belong to the primary landmark population. The prediction cohort requires database coverage and survival for 90 days after the first systolic blood pressure reading greater than 140 mmHg. A separate sample of 31,390 patients with any recorded data after that reading was used to analyze death as a competing event. Denominators for all analysis samples are provided in Table S9. (B) Timing of study measures. Covariates were assessed over the 365 days preceding index. Contact trajectory classes were derived from quarters 1–7 (days 91–727), and the contact-gap measure covered days 0–730. Cardiovascular hospitalization and death as a competing event were assessed for up to 730 days after the day-730 landmark. The prediction window began at the first systolic blood pressure reading greater than 140 mmHg (median, day 22) and extended for 90 days. Contact measures reflect records within a single health system. BP, blood pressure; CV, cardiovascular.

### Prediction of no recorded blood pressure measurement

For each patient, predictions were generated at the first systolic blood pressure reading greater than 140 mmHg on or after the index date. Patients were included in the prediction cohort if database coverage extended at least 90 days beyond the prediction date and they survived through day 90. The outcome was the absence of a recorded blood pressure measurement within 90 days of the prediction date. Because measurements obtained outside the observed health system are not captured, this outcome reflects absent documentation rather than necessarily indicating a missed clinical reassessment. The prediction trigger was based on systolic blood pressure alone. Among qualifying readings, 26.7% were accompanied by a diastolic pressure of at least 90 mmHg. In addition, 9.7% of patients had an earlier post-index reading with a diastolic pressure of at least 90 mmHg and a systolic pressure of 140 mmHg or lower; these readings did not trigger the prediction window. Guidelines recommend reassessment approximately 1 month after treatment initiation or adjustment and within 3 to 6 months when blood pressure is managed with lifestyle modification alone [5]. We selected a 90-day prediction window to accommodate the recommended early reassessment while allowing additional time for scheduling. In this cohort, the median time to the next recorded blood pressure measurement was 35 days (interquartile range [IQR], 14–91). Alternative outcome definitions using 30-, 180-, 270-, and 365-day windows, as well as analyses treating death before any subsequent blood pressure measurement as an outcome or as a competing event, are described in the Supplementary Methods.

The model incorporated 62 features derived exclusively from information available at the prediction time: demographic characteristics, insurance, Elixhauser comorbidities, laboratory values, body mass index and associated missingness indicators, blood pressure level and longitudinal variability and trend, medication count, prior encounters, time since the last visit, and visit regularity.[24] Contact trajectory class was excluded because it was measured after the prediction date for most patients. Patients were randomly assigned, with stratification by outcome, to training (60%), validation (20%), and test (20%) sets; thus, all model evaluation was internal. We compared a two-variable logistic regression model using time since the last visit and prior encounters, an L2-penalized logistic regression model, and a gradient-boosted tree model [25]. Hyperparameters were tuned using five-fold cross-validation within the training set. The selected models were recalibrated using isotonic regression on the validation set and evaluated once on the held-out test set.

We assessed discrimination using the area under the receiver operating characteristic curve (AUROC) and the area under the precision–recall curve (AUPRC) [26]. Calibration was evaluated using the Brier score and calibration intercept and slope [27]. Paired AUROC differences were estimated, and 95% confidence intervals were obtained using 1,000 bootstrap samples. Shapley values were used to characterize feature contributions to model predictions [28]. To assess the robustness of the results to the random data split, we also performed nested five-fold cross-validation to generate out-of-fold predictions for every patient (Supplementary Methods). Reporting followed the Transparent Reporting of a Multivariable Prediction Model for Individual Prognosis or Diagnosis (TRIPOD) guidelines [29].

Because outreach costs depend substantially on the number of patients a clinic can contact, we evaluated model performance at fixed outreach capacities. Specifically, we flagged the 5%, 10%, 20%, or 30% of patients in the test set with the highest predicted risk. At each capacity level, we calculated the proportion of patients without a recorded blood pressure measurement who were flagged (sensitivity), the proportion of flagged patients who had no recorded measurement (positive predictive value), and the number of outreach contacts required per patient identified. We estimated 95% confidence intervals using bootstrap resampling. Decision curve analysis [7] was also performed to estimate net benefit across risk thresholds, with 95% bootstrap confidence bands.

For the equity audit, we flagged the 20% of patients with the highest out-of-fold predicted risk [9]. Within racial and ethnic, insurance, sex, and age groups, we assessed AUROC, calibration (observed-to-predicted ratio and calibration slope), the proportion flagged, true-positive and false-positive rates, and positive predictive value. We calculated bootstrap 95% confidence intervals for each measure and for differences in true-positive rates relative to a reference group [30]. Because a model can be calibrated within each group yet achieve different detection rates when outcome prevalence varies across groups [10], we additionally evaluated group-specific decision thresholds designed to equalize detection rates across groups.

### Landmark and follow-up for the association analysis

The association analysis began with the at-index cohort, comprising patients whose database coverage extended through day 730 after index. Because both contact measures were derived from the first two years after index, cardiovascular outcomes were ascertained from a landmark at day 730 [18, 19]. The primary landmark population excluded patients with a recorded cardiovascular hospitalization before index or during the interval from index through day 730. This requirement relied on information obtained after the landmark and excluded 1,476 patients whose survival to day 730 was known but who had no subsequent record. To assess the potential effect of this selection, we repeated the cause-specific analyses in a day-730-information sample, in which survival at the landmark was confirmed solely by hospital or emergency department contact during days 366–730 (**Figure 1A**; **Table 4**). This alternative sample was also subject to selection because inclusion required recorded contact during the second year after index.

Follow-up ended at the earliest of the end of database coverage, day 1,460 after index, 365 days after the last hospital or emergency department contact, or death. The same follow-up end date was used for cardiovascular hospitalization and death. Accordingly, the results apply to patients who were alive, free of prior cardiovascular hospitalization, and eligible for follow-up based on recorded data two years after index. **Figure 1** illustrates cohort selection and the timing of covariate measurement, contact assessment, and outcome follow-up.

### Contact measures

Contact was measured in consecutive 91-day quarters as the number of unique days on which any encounter, blood pressure measurement, or antihypertensive prescription order was recorded. Multiple types of contact on the same day counted as a single contact day. This measure captured the frequency of recorded contact rather than the number of records generated and did not assume that more contact represented better care. Contact trajectories were derived from quarters 1–7 (days 91–727), ensuring that all trajectory measurements preceded the day-730 landmark. Let *U_iq_* denote the number of unique contact days for patient *i* in quarter *q*. The trajectory models used the transformed contact measure

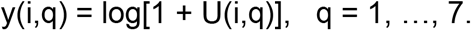

Group-based trajectory models [13] were fitted with the lcmm package [31] in the landmark population, using only pre-landmark contact data. Solutions containing one to six classes were compared using model selection criteria, without reference to outcome data [32]. Because the information criterion continued to improve as the number of classes increased, we selected the smallest adequate solution: a four-class model. This was the fewest-class solution that distinguished rising from declining contact trajectories while maintaining adequate classification quality across all classes (**Table S2**; Supplementary Methods). Each patient was assigned to the class with the highest posterior probability of membership.

To account for uncertainty in class assignment, we conducted a sensitivity analysis using each patient’s posterior class-membership probabilities in place of the assigned class, allowing patients to contribute proportionally to each class according to their estimated membership probabilities. A five-class solution, which further divided the sustained-contact trajectory into moderate- and high-contact classes, was examined in a second sensitivity analysis.

For the contact-gap measure, let *d*_i(1)_ <⋯<*d*_i(m)_ denote the ordered days from day 0 through day 730 on which patient i had a recorded blood pressure measurement or hospital admission, with the index reading defined as day 0. The longest interval without recorded contact was defined as

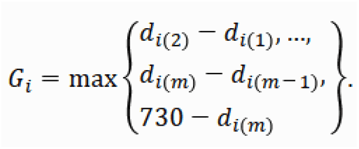

This definition included the interval from the last recorded contact to the landmark, thereby allowing a gap to be measured without requiring a subsequent return. The longest gap was modeled as a continuous exposure using a restricted cubic spline [33], with a 90-day interval as the reference. Gaps exceeding 180, 270, and 365 days were examined in secondary analyses because no universally accepted threshold defines a clinically meaningful gap in care [11]. A gap represented an absence of recorded contact within the observed health system, not confirmed disengagement from health care.

### Outcome and covariates

For the association analysis, the outcome was the first hospitalization after the day-730 landmark with myocardial infarction, stroke, or heart failure recorded as the principal diagnosis. We used the principal diagnosis to improve the specificity of the hospitalization endpoint, particularly because chronic heart failure may be coded during admissions for unrelated conditions [34]. Accordingly, the outcome represented cardiovascular hospitalization rather than incident cardiovascular disease. Death from any cause before cardiovascular hospitalization was treated as the competing event because cause-specific mortality information was unavailable.

Covariates included age, sex, race and ethnicity as recorded in the EHR, insurance, systolic and diastolic blood pressure, estimated glomerular filtration rate (eGFR), glycated hemoglobin, body mass index, Elixhauser comorbidity score [35, 36], prior emergency department visits, prior inpatient admissions, total prior encounters, antihypertensive medication count, and characteristics of the index episode. All covariates were derived from information available on or before the index date. Blood pressure measurements were taken from the index date, whereas laboratory values and body mass index were taken from the measurements closest to index within the preceding 365 days. eGFR was calculated using the 2021 CKD-EPI equation without a race coefficient. Age, systolic blood pressure, eGFR, body mass index, Elixhauser score, prior encounters, prior inpatient admissions, and antihypertensive medication count were modeled using restricted cubic splines [33].

Diastolic blood pressure and glycated hemoglobin were modeled as linear terms. Missing values for eGFR (5.6%), glycated hemoglobin (60.9%), and body mass index (30.9%) were handled using multiple imputation by chained equations, with 30 imputed datasets [37, 38]. Estimates were combined using Rubin’s rules [39]. Sensitivity analyses used missing-indicator methods [40], complete-case analysis, and delta-adjusted imputation to assess the robustness of findings to assumptions about missing data [41] (Supplementary Methods).

### Association analyses

Cumulative incidence was estimated using the Aalen–Johansen estimator and compared across groups using Gray’s test [42]. Cause-specific Cox proportional hazards models were used to estimate associations with cause-specific event rates, and Fine–Gray subdistribution hazards models were used to assess associations with cumulative incidence in the presence of competing mortality [43, 44]. The proportional hazards assumption was evaluated using scaled Schoenfeld residuals [45]. Doubly robust estimates combined stabilized inverse probability weights, truncated at the 1^st^ and 99^th^ percentiles, with covariate adjustment [46–48]. To account for potentially informative loss to follow-up, we additionally applied inverse probability-of-censoring weights. The probability of remaining under follow-up was estimated at 91-day intervals using pooled logistic regression incorporating covariates and both contact measures. The resulting stabilized weights were truncated at the 1^st^ and 99^th^ percentiles [46]. To assess whether contact patterns provided information beyond the covariates, we compared a covariate-only model with models additionally including the contact-gap measure or trajectory class. Incremental information was assessed using pooled Wald tests and the change in Uno’s C-statistic at 730 days [49]. All models were fitted separately in each imputed dataset, and estimates were pooled across imputations. Sensitivity analyses are summarized in Table S3. We also report the minimum detectable hazard ratio at 80% power [50].

All tests were two-sided, with a significance threshold of α = 0.05. No adjustment was made for multiple comparisons. Analyses of component outcomes were considered exploratory and comprised 12 comparisons: one gap contrast (greater than 365 days versus the reference) and three trajectory-class contrasts, each evaluated for myocardial infarction, stroke, and heart failure hospitalization. Software details and a supplementary analysis of comorbidity patterns are provided in the Supplementary Methods.

## RESULTS

### Study population

Among 50,946 adults with a hypertension diagnosis and a subsequent blood pressure measurement, 48,628 met the index-cohort eligibility criteria. The prediction cohort comprised 30,942 patients, while the at-index cohort comprised 37,183 patients, of whom 15,200 were included in the primary landmark population (**Figure 1A**). The largest exclusion from the landmark population comprised 14,361 patients whose survival to day 730 could not be confirmed because they had neither a hospital or emergency department contact during days 366–730 nor a subsequent record. Consequently, the landmark analysis excluded a substantial number of patients without evidence of continued contact within the observed health system. An additional 1,476 patients were known to be alive at day 730 but had no subsequent record; they were excluded from the primary landmark population because inclusion required a record after day 730. Among the 32,101 patients with a systolic blood pressure reading greater than 140 mmHg, 711 had no recorded data after the qualifying reading, 254 had insufficient database coverage for the 90-day prediction horizon, and 194 died within 90 days. These patients were excluded from the prediction cohort, leaving 30,942 patients for analysis (**Table S9**).

### Prediction of no recorded blood pressure measurement

Among the 30,942 patients in the prediction cohort, 10,899 (35.2%) had no blood pressure measurement recorded in the health system within 90 days of the qualifying elevated reading. The median interval between index and prediction was 22 days. Patients without a recorded measurement were broadly similar to those with a subsequent measurement in age, sex, race and ethnicity, insurance, blood pressure measurements at and before the qualifying reading, laboratory values, and comorbidity burden; all standardized differences were ≤0.13. However, patients without a recorded measurement had fewer prior encounters (median, 4 vs 8), a longer interval since their last visit (median, 55 vs 22 days), and more frequently had insufficient prior readings to estimate blood pressure variability (45.1% vs 23.7%) (**Table 1**; **Table S4**).

**Table 1.** Characteristics of the prediction sample at the first elevated reading, by whether a blood pressure measurement was recorded within 90 days.

| Characteristic | Overall | No recorded BP in 90 days | BP recorded in 90 days | SMD |
| --- | --- | --- | --- | --- |
| Patients, n | 30,942 | 10,899 | 20,043 |  |
| Age, years, mean (SD) | 66.6 (13.6) | 67.2 (13.7) | 66.3 (13.6) | 0.07 |
| Female, n (%) | 16,032 (51.8) | 5,365 (49.2) | 10,667 (53.2) | 0.08 |
| **Race and ethnicity, n (%)** |  |  |  |  |
| White | 20,344 (65.7) | 7,283 (66.8) | 13,061 (65.2) | 0.03 |
| Black | 5,792 (18.7) | 1,845 (16.9) | 3,947 (19.7) | 0.07 |
| Hispanic | 1,710 (5.5) | 524 (4.8) | 1,186 (5.9) | 0.05 |
| Asian | 1,002 (3.2) | 349 (3.2) | 653 (3.3) | 0.00 |
| Other or unknown | 2,094 (6.8) | 898 (8.2) | 1,196 (6.0) | 0.09 |
| **Insurance, n (%)** |  |  |  |  |
| Medicare | 14,225 (46.0) | 5,133 (47.1) | 9,092 (45.4) | 0.03 |
| Medicaid | 1,926 (6.2) | 621 (5.7) | 1,305 (6.5) | 0.03 |
| Other | 14,791 (47.8) | 5,145 (47.2) | 9,646 (48.1) | 0.02 |
| **Clinical** |  |  |  |  |
| Systolic BP at qualifying reading, mmHg, mean (SD) | 152.4 (11.2) | 152.7 (11.1) | 152.2 (11.3) | 0.05 |
| Diastolic BP at qualifying reading, mmHg, mean (SD) | 80.7 (13.4) | 80.7 (13.3) | 80.8 (13.5) | 0.01 |
| Systolic BP, earlier readings, mmHg, mean (SD) | 132.2 (13.0) | 132.1 (13.2) | 132.3 (12.9) | 0.02 |
| Diastolic BP, earlier readings, mmHg, mean (SD) | 75.2 (9.9) | 75.1 (10.3) | 75.2 (9.7) | 0.00 |
| eGFR, mL/min/1.73 m<sup>2</sup>, median (IQR) | 77 (56–94) | 77 (56–93) | 77 (56–94) | 0.01 |
| HbA1c, %, median (IQR) | 6.1 (5.7–7.1) | 6.1 (5.6–7.0) | 6.1 (5.7–7.1) | 0.04 |
| BMI, kg/m<sup>2</sup>, mean (SD) | 30.2 (7.1) | 30.1 (7.1) | 30.3 (7.1) | 0.03 |
| Elixhauser score, median (IQR) | 5 (0–11) | 5 (0–10) | 5 (0–12) | 0.13 |
| Antihypertensive drugs, median (IQR) | 2 (0–3) | 2 (0–3) | 2 (0–3) | 0.02 |
| **Contact history before prediction** |  |  |  |  |
| Prior encounters, median (IQR) | 6 (3–13) | 4 (2–8) | 8 (4–16) | 0.45 |
| Any prior emergency visit, n (%) | 4,277 (13.8) | 1,301 (11.9) | 2,976 (14.8) | 0.09 |
| Days since last visit, median (IQR) | 30 (13–82) | 55 (24–133) | 22 (10–55) | 0.33 |
| **Missing values, n (%)** |  |  |  |  |
| eGFR | 1,711 (5.5) | 858 (7.9) | 853 (4.3) | 0.15 |
| HbA1c | 18,222 (58.9) | 6,744 (61.9) | 11,478 (57.3) | 0.09 |
| BMI | 5,942 (19.2) | 2,940 (27.0) | 3,002 (15.0) | 0.30 |
| Days since last visit | 3,200 (10.3) | 1,628 (14.9) | 1,572 (7.8) | 0.22 |
| Systolic BP variability (fewer than two readings) | 9,668 (31.2) | 4,914 (45.1) | 4,754 (23.7) | 0.46 |

Note: Characteristics were assessed at the prediction date, defined as the first systolic blood pressure reading greater than 140 mmHg on or after index. The qualifying reading was the measurement that triggered prediction and therefore had a systolic pressure greater than 140 mmHg in all patients. Earlier blood pressure measurements were summarized as the mean of all readings before the prediction date; 5,928 patients (19.2%) had no earlier readings. Laboratory values, body mass index, and antihypertensive medication count were assessed over the 365 days up to and including the prediction date; contact history was derived from earlier dates, and comorbidities were identified from diagnoses recorded on or before index. No recorded blood pressure measurement was defined as the absence of a blood pressure measurement documented within the health system during the subsequent 90 days. Summaries were calculated using observed values. Characteristics by training, validation, and test set are provided in Table S4. BMI, body mass index; BP, blood pressure; eGFR, estimated glomerular filtration rate; HbA1c, glycated hemoglobin; IQR, interquartile range; SD, standard deviation; SMD, standardized mean difference between patients with and without a recorded measurement.

In the test set (n = 6,189; 35.3% without a recorded measurement), the gradient-boosted tree model achieved an AUROC of 0.740 (95% CI, 0.727–0.753) and an AUPRC of 0.568 (95% CI, 0.546–0.593). Discrimination was higher than that of penalized logistic regression (AUROC difference, 0.042; 95% CI, 0.033–0.052) and the two-variable logistic regression model (AUROC difference, 0.054; 95% CI, 0.044–0.063) (Table 2A). Nested cross-validation across all 30,942 patients yielded similar performance, with an AUROC of 0.746 (95% CI, 0.741–0.751) and an AUPRC of 0.578 (95% CI, 0.569–0.589). Model predictions were reasonably well calibrated: the test-set Brier score was 0.193, compared with 0.228 for the prevalence-only benchmark, and the calibration slope was 0.91 in the test set and 0.98 across cross-validation folds (**Figure S1**).

**Table 2.** Performance of models predicting no recorded blood pressure measurement within 90 days of an elevated reading.

Discrimination and calibration (test set, n = 6,189; 2,185 with no recorded BP)
| Model | AUROC (95% CI) | AUPRC (95% CI) | Brier score | Calibration intercept / slope | AUROC difference, boosted minus model (95% CI) |
| --- | --- | --- | --- | --- | --- |
| Prevalence only | 0.500 | 0.353 | 0.228 | — | — |
| Two-variable logistic (days since last visit, prior encounters) | 0.687 (0.672–0.700) | 0.508 (0.486–0.531) | 0.206 | –0.01 / 0.93 | +0.054 (0.044 to 0.063) |
| Penalized logistic regression (62 features) | 0.698 (0.685–0.712) | 0.515 (0.493–0.539) | 0.204 | –0.01 / 0.92 | +0.042 (0.033 to 0.052) |
| Gradient-boosted trees (62 features) | 0.740 (0.727–0.753) | 0.568 (0.546–0.593) | 0.193 | –0.01 / 0.91 | — |
| Gradient-boosted trees, nested cross-validation (all 30,942) | 0.746 (0.741–0.751) | 0.578 (0.569–0.589) | — | 0.00 / 0.98 | — |

**B. Outreach at fixed capacity: flagging the patients with the highest predicted risk (test set)**
| Patients flagged | Flagged, n | Selected patients with no recorded BP, n | Share of patients with no recorded BP found, % (95% CI) | Flagged patients with no recorded BP (PPV), % (95% CI) | Patients selected per outcome-positive patient (95% CI) | Sensitivity, %: penalized logistic / two-variable |
| --- | --- | --- | --- | --- | --- | --- |
| Top 5% | 309 | 205 | 9.4 (8.7–10.2) | 66.3 (61.2–71.8) | 1.5 (1.4–1.6) | 7.9 / 8.0 |
| Top 10% | 619 | 383 | 17.5 (16.5–18.8) | 61.9 (58.5–66.2) | 1.6 (1.5–1.7) | 16.4 / 16.4 |
| Top 20% | 1,238 | 748 | 34.2 (32.7–35.7) | 60.4 (57.4–63.2) | 1.7 (1.6–1.7) | 31.9 / 32.0 |
| Top 30% | 1,857 | 1,076 | 49.2 (47.6–50.9) | 57.9 (55.6–60.3) | 1.7 (1.7–1.8) | 46.4 / 47.1 |

Note: The outcome was the absence of a blood pressure measurement recorded within the health system during the 90 days after the first systolic blood pressure reading greater than 140 mmHg, among patients who survived and had database coverage throughout the 90-day period. Brier scores and calibration measures were calculated using isotonic-recalibrated predictions. Confidence intervals were estimated from 1,000 bootstrap samples, except in nested cross-validation, for which 500 bootstrap samples were used. In panel B, patients are ranked by predicted risk from the gradient-boosted tree model. Under random selection, 35.3% of flagged patients would have no recorded measurement, corresponding to approximately 2.8 outreach contacts per patient identified. AUPRC, area under the precision–recall curve; AUROC, area under the receiver operating characteristic curve; PPV, positive predictive value.

The interval since the last visit was the largest contributor to predictions, accounting for 30% of the total Shapley value contribution, followed by the average interval between visits, prior encounter frequency, and systolic blood pressure variability. Longer intervals since the last visit and fewer prior encounters were associated with higher predicted risk.

### Outreach yield and decision curve analysis

Flagging the 20% of patients with the highest predicted risk identified 34.2% (95% CI, 32.7–35.7) of patients who subsequently had no recorded blood pressure measurement. Among flagged patients, 60.4% (95% CI, 57.4–63.2) had no recorded measurement, corresponding to approximately 1.7 outreach contacts per patient identified, compared with 2.8 contacts under random selection (**Table 2B; Figure 2A**). Across selection thresholds ranging from the top 5% to the top 30% of predicted risk, positive predictive value remained between 58% and 66%, suggesting that outreach capacity could be adjusted within this range without substantial variation in yield. The full model identified more patients without a recorded measurement than either simpler model at every capacity level. In decision curve analysis, the model yielded greater net benefit than the strategies of contacting everyone or no one across the evaluated risk thresholds of 0.05–0.70. The lower bound of the 95% confidence interval for net benefit also exceeded both reference strategies over thresholds of 0.15–0.58. Results were consistent across alternative prediction horizons of 30–365 days, for which AUROC values ranged from 0.737 to 0.791 (**Table S5**). Death before a subsequent recorded blood pressure measurement was uncommon across all predicted-risk fifths, ranging from 0.1% to 0.4%.

**Figure 2.**
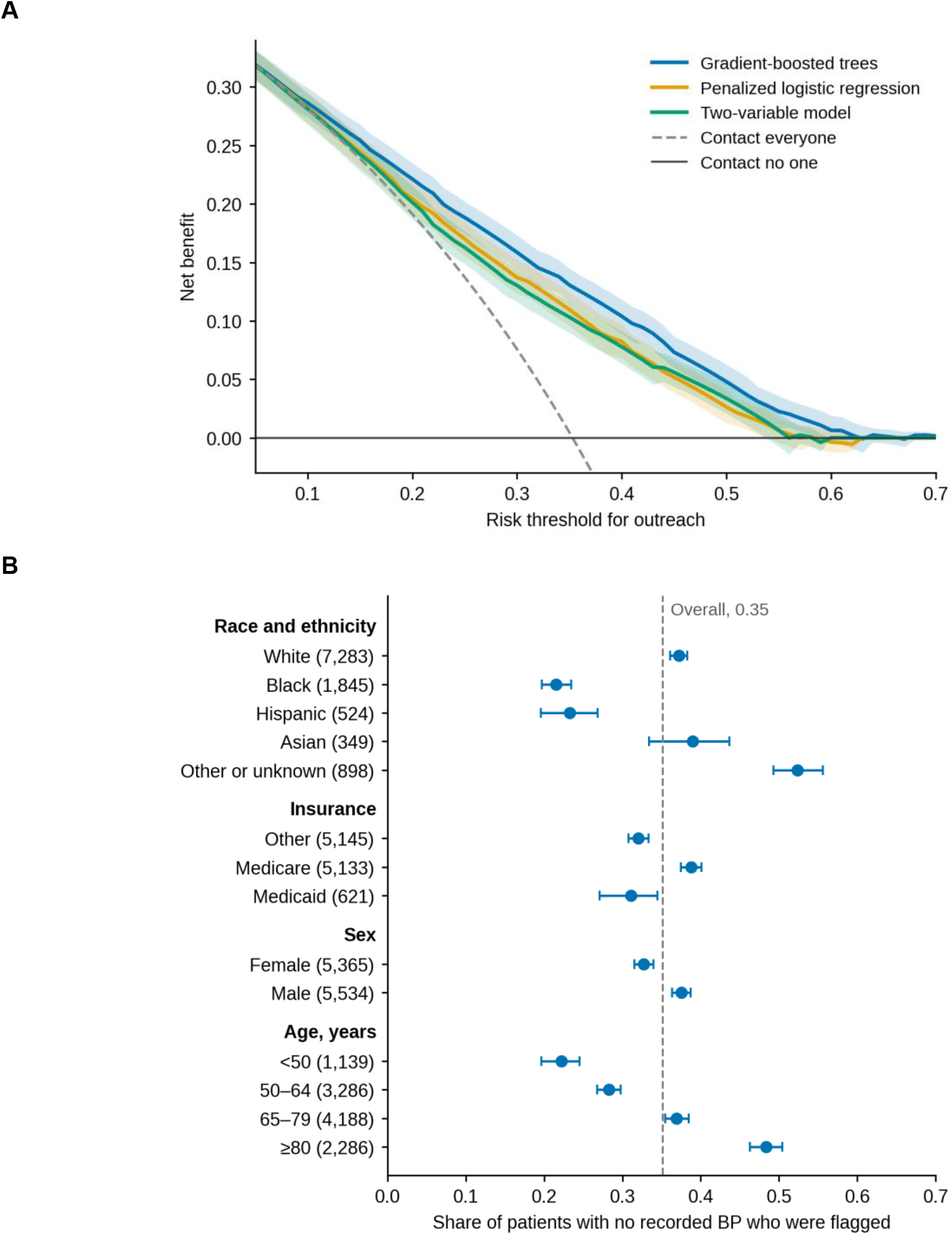
Clinical utility and subgroup performance of the model predicting the absence of a recorded blood pressure measurement within 90 days. (A) Decision curves in the test set (n = 6,189; 2,185 patients without a recorded measurement) comparing gradient-boosted trees, penalized logistic regression, and the two-variable logistic regression model with the strategies of contacting everyone or no one. Shaded bands represent bootstrap 95% confidence intervals based on 1,000 resamples. (B) Proportion of patients without a recorded measurement identified at an overall selection rate of 20%, based on the highest predicted risks from out-of-fold predictions (n = 30,942), stratified by subgroup. Error bars represent bootstrap 95% confidence intervals. The dashed line indicates the overall detection rate (0.35); numbers in parentheses indicate the number of patients without a recorded measurement in each group.

### Performance by groups

**Table 3** presents the full set of performance measures across subgroups. Discrimination was similar across groups, with AUROC values ranging from 0.71 to 0.78. Mean predicted risk was also consistent with observed outcome prevalence across groups, with observed-to-predicted ratios ranging from 0.96 to 1.03. Calibration slopes ranged from 0.87 to 1.09; their 95% confidence intervals included 1 in all groups except Black patients (95% CI, 0.82–0.98) and patients younger than 50 years (95% CI, 0.78–0.97). Despite broadly similar discrimination and average calibration, outcome detection differed substantially across groups (**Figure 2B**). When the 20% of patients with the highest predicted risk were flagged, the model identified 37.2% of White patients without a recorded measurement, compared with 21.5% of Black patients (difference, −0.16; 95% CI, −0.18 to −0.14) and 23.3% of Hispanic patients (difference, −0.14; 95% CI, −0.18 to −0.10). The corresponding detection rate was 52.3% among patients categorized as having other or unknown race. Detection also varied by age: it was 22.2% among patients younger than 50 years (difference, −0.15; 95% CI, −0.18 to −0.12), 28.3% among those aged 50–64 years, and 48.4% among those aged 80 years or older. The difference for Medicaid beneficiaries was smaller, with a detection rate of 31.1% (difference, −0.01; 95% CI, −0.05 to 0.03). Differences in detection by race and ethnicity exceeded the corresponding differences in outcome prevalence. The proportion of patients without a recorded measurement was 31.9% among Black patients and 30.6% among Hispanic patients, compared with 35.8% among White patients. However, the proportion selected for outreach was also lower among Black and Hispanic patients (12.5% and 13.4%, respectively) than among White patients (21.3%). Thus, similar overall discrimination and average calibration did not translate into comparable detection at a common outreach capacity. In an additional analysis, we selected separate thresholds for each racial and ethnic group to achieve a common sensitivity equal to the overall detection rate of 35.2% (**Table S6**). This approach maintained the overall proportion of patients flagged at 20.0% but increased the false-positive rate among Black patients from 0.08 to 0.13 and among Hispanic patients from 0.09 to 0.13, while positive predictive values remained similar.

**Table 3.** Model performance by subgroup with the 20% of patients at highest predicted risk flagged (out-of-fold predictions; n = 30,942; 10,899 with no recorded BP; estimates with 95% CIs)

| Group | n | No recorded BP, % | AUROC | Observed / predicted | Calibration slope | Flagged | TPR | FPR | PPV | TPR difference (95% CI) |
| --- | --- | --- | --- | --- | --- | --- | --- | --- | --- | --- |
| **Race and ethnicity** |  |  |  |  |  |  |  |  |  |  |
| White (reference) | 20,344 | 35.8 | 0.749<br>(0.742–0.755) | 1.00<br>(0.99–1.02) | 0.99<br>(0.96–1.03) | 0.213<br>(0.208–0.219) | 0.372<br>(0.361–0.383) | 0.125<br>(0.119–0.130) | 0.625<br>(0.612–0.638) | — |
| Black | 5,792 | 31.9 | 0.720<br>(0.706–0.734) | 0.98<br>(0.95–1.02) | 0.90<br>(0.82–0.98) | 0.125<br>(0.117–0.133) | 0.215<br>(0.197–0.235) | 0.083<br>(0.073–0.091) | 0.549<br>(0.512–0.588) | –0.16 (–0.18 to –0.14) |
| Hispanic | 1,710 | 30.6 | 0.731<br>(0.705–0.758) | 0.96<br>(0.90–1.03) | 0.89<br>(0.72–1.05) | 0.134<br>(0.118–0.148) | 0.233<br>(0.196–0.268) | 0.090<br>(0.073–0.106) | 0.533<br>(0.465–0.601) | –0.14 (–0.18 to –0.10) |
| Asian | 1,002 | 34.8 | 0.777<br>(0.746–0.806) | 1.03<br>(0.95–1.11) | 1.09<br>(0.91–1.29) | 0.198<br>(0.172–0.223) | 0.390<br>(0.334–0.436) | 0.095<br>(0.072–0.119) | 0.687<br>(0.615–0.753) | +0.02 (–0.04 to +0.06) |
| Other or unknown | 2,094 | 42.9 | 0.752<br>(0.733–0.773) | 1.03<br>(0.99–1.08) | 1.01<br>(0.90–1.13) | 0.335<br>(0.317–0.357) | 0.523<br>(0.493–0.556) | 0.194<br>(0.174–0.215) | 0.670<br>(0.636–0.703) | +0.15 (+0.12 to +0.18) |
| **Insurance** |  |  |  |  |  |  |  |  |  |  |
| Other (reference) | 14,791 | 34.8 | 0.742<br>(0.733–0.750) | 1.00<br>(0.98–1.02) | 0.97<br>(0.92–1.02) | 0.183<br>(0.177–0.190) | 0.321<br>(0.308–0.334) | 0.110<br>(0.104–0.116) | 0.608<br>(0.590–0.626) | — |
| Medicare | 14,225 | 36.1 | 0.749<br>(0.740–0.757) | 1.00<br>(0.98–1.02) | 0.98<br>(0.94–1.02) | 0.224<br>(0.218–0.230) | 0.388<br>(0.375–0.401) | 0.131<br>(0.125–0.138) | 0.625<br>(0.608–0.641) | +0.07 (+0.05 to +0.09) |
| Medicaid | 1,926 | 32.2 | 0.752<br>(0.728–0.774) | 0.98<br>(0.92–1.03) | 1.03<br>(0.89–1.16) | 0.151<br>(0.134–0.165) | 0.311<br>(0.271–0.345) | 0.074<br>(0.060–0.089) | 0.666<br>(0.608–0.722) | –0.01 (–0.05 to +0.03) |
| **Sex** |  |  |  |  |  |  |  |  |  |  |
| Female (reference) | 16,032 | 33.5 | 0.745<br>(0.737–0.754) | 0.99<br>(0.97–1.01) | 0.97<br>(0.93–1.02) | 0.178<br>(0.173–0.184) | 0.327<br>(0.316–0.340) | 0.103<br>(0.097–0.108) | 0.615<br>(0.600–0.635) | — |
| Male | 14,910 | 37.1 | 0.745<br>(0.738–0.753) | 1.01<br>(0.99–1.03) | 0.99<br>(0.94–1.03) | 0.224<br>(0.217–0.230) | 0.375<br>(0.363–0.387) | 0.134<br>(0.127–0.141) | 0.623<br>(0.608–0.639) | +0.05 (+0.03 to +0.07) |
| **Age** |  |  |  |  |  |  |  |  |  |  |
| 65–79 years (reference) | 11,921 | 35.1 | 0.752<br>(0.743–0.762) | 1.00<br>(0.97–1.02) | 0.98<br>(0.93–1.04) | 0.207<br>(0.200–0.215) | 0.369<br>(0.355–0.385) | 0.120<br>(0.113–0.127) | 0.625<br>(0.609–0.644) | — |
| <50 years | 3,372 | 33.8 | 0.710<br>(0.691–0.727) | 1.00<br>(0.96–1.05) | 0.87<br>(0.78–0.97) | 0.133<br>(0.121–0.143) | 0.222<br>(0.196–0.245) | 0.087<br>(0.076–0.099) | 0.565<br>(0.521–0.610) | –0.15 (–0.17 to –0.12) |
| 50–64 years | 9,729 | 33.8 | 0.736<br>(0.727–0.746) | 0.99<br>(0.97–1.02) | 0.97<br>(0.91–1.02) | 0.163<br>(0.156–0.171) | 0.283<br>(0.268–0.298) | 0.102<br>(0.094–0.109) | 0.587<br>(0.560–0.611) | –0.09 (–0.11 to –0.06) |
| ≥80 years | 5,920 | 38.6 | 0.764<br>(0.751–0.775) | 1.01<br>(0.98–1.04) | 1.04<br>(0.97–1.10) | 0.284<br>(0.273–0.295) | 0.484<br>(0.463–0.504) | 0.159<br>(0.147–0.170) | 0.657<br>(0.635–0.680) | +0.12 (+0.09 to +0.14) |

Note: Estimates are presented with bootstrap 95% confidence intervals based on 500 resamples. Differences in the true-positive rate (TPR) are calculated relative to the reference group within each demographic or clinical dimension. Calibration was assessed using isotonic recalibration fitted on the other four folds. No recorded BP was defined as the absence of a blood pressure measurement documented within the health system during the subsequent 90 days. Race and ethnicity were incompletely recorded in the database. AUROC, area under the receiver operating characteristic curve; FPR, false-positive rate; PPV, positive predictive value; TPR, true-positive rate (proportion of patients without a recorded measurement who were flagged).

### Contact patterns and cardiovascular hospitalization

Within 730 days after the landmark, 664 patients (4.4%) experienced a cardiovascular hospitalization, whereas 1,077 (7.1%) died before experiencing the outcome. An additional 3,556 patients (23.4%) were censored before the end of follow-up, at a median of 443 days after the landmark. Of these, 3,028 were censored when death ascertainment ended and 528 when database coverage ended. Early censoring was modestly more frequent among patients classified as having Declining, Rising, or Sustained Contact than among those with Minimal Contact (odds ratios, 1.09–1.35) and among patients with a contact gap than among those without one (odds ratio, 1.11; 95% CI, 1.01–1.23). Inverse probability weighting for censoring did not materially change the estimates for the association between a contact gap and cardiovascular hospitalization (hazard ratio [HR], 0.86; 95% CI, 0.71–1.05) or between Sustained Contact and the outcome (HR, 1.39; 95% CI, 1.04–1.84; **Table S3**).

Four distinct contact patterns were identified (**Figure 3A**): Minimal Contact (49.8% of the landmark population), Declining Contact (22.5%), Rising Contact (17.6%), and Sustained Contact (10.1%). The Bayesian information criterion (BIC) decreased with each additional class through the six-class solution (**Table S2**); therefore, BIC alone did not identify a definitive number of classes. The final four-class solution was selected according to the prespecified classification criteria described in the Methods. It was the smallest solution that distinguished rising from declining contact while maintaining adequate classification performance across all classes, with average posterior probabilities ranging from 0.74 to 0.92 and odds of correct classification of at least 9.9. The five-class solution subdivided Sustained Contact into moderate- and high-contact groups, whereas the six-class solution introduced two additional classes, each comprising less than 4% of the population, and further subdivided Rising Contact without identifying a clearly distinct additional pattern. Despite these classification metrics, uncertainty remained for a subset of patients: 23.0% had a maximum posterior class-membership probability below 0.70. Incorporating posterior probabilities into the outcome model, rather than relying solely on assigned classes, yielded a similar estimate for Sustained Contact (HR, 1.42; 95% CI, 1.05–1.93, versus HR, 1.40 in the assigned-class analysis; Table S3). The five-class solution likewise produced a similar estimate for Sustained Moderate Contact (HR, 1.41; 95% CI, 1.11–1.79). Differences between classes were more pronounced in clinical characteristics and prior health care utilization than in demographic characteristics (**Table S7**). Median Elixhauser comorbidity scores ranged from 4 in the Minimal Contact class to 7 in the Sustained Contact class, with a maximum standardized difference of 0.49. Median numbers of prior health care encounters ranged from 3 to 9, with a maximum standardized difference of 0.71. In contrast, standardized differences for age, blood pressure, laboratory measures, sex, race, and insurance status were 0.17 or lower. The availability of clinical measurements also varied across classes. For example, body mass index was missing for 38.8% of patients in the Minimal Contact class, compared with 13.2% in the Sustained Contact class.

**Figure 3.**
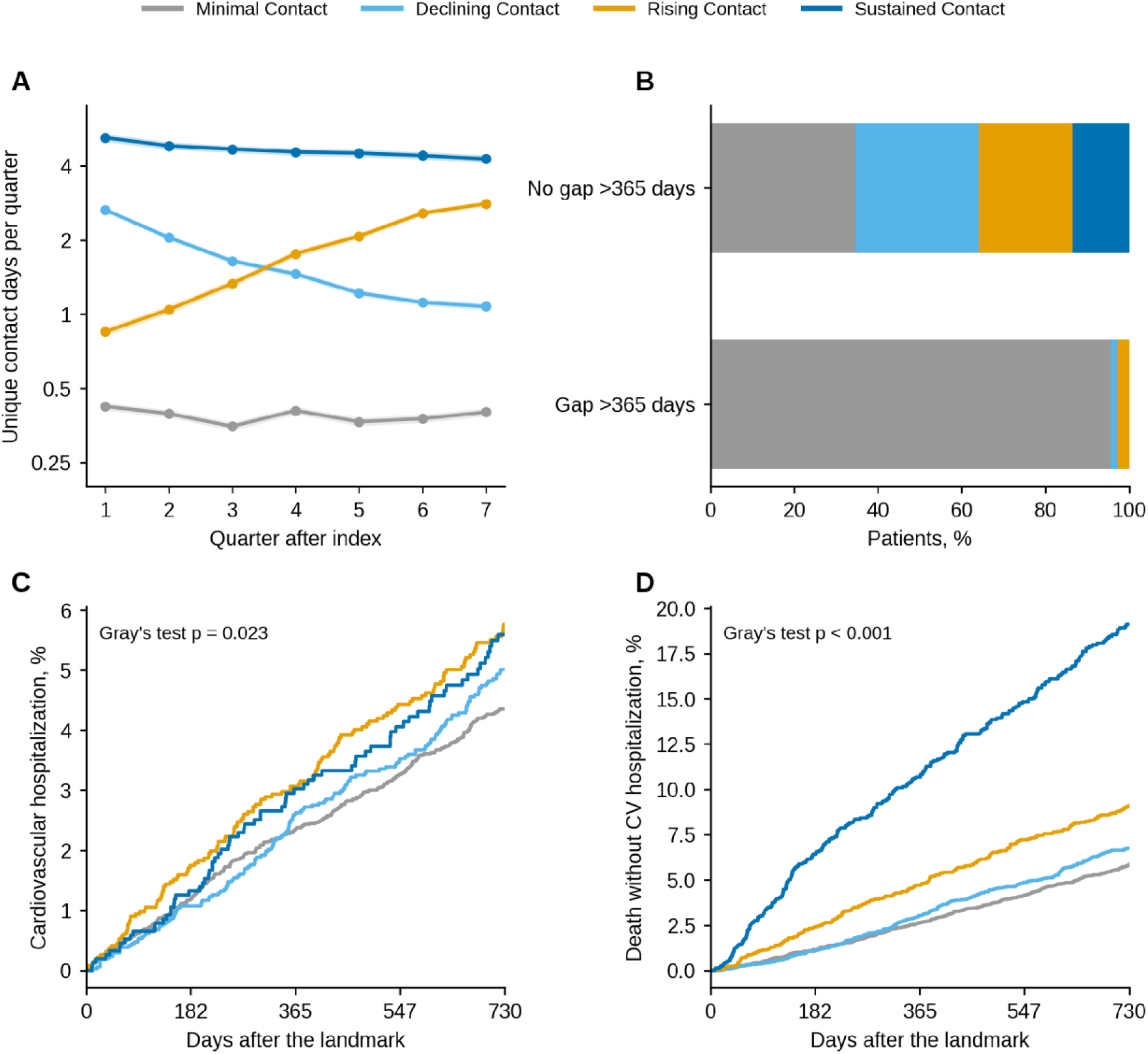
Contact patterns and outcomes in the landmark population (n = 15,200). (A) Unique contact days per quarter (days with an encounter, blood pressure reading, or prescription order) by trajectory class: backtransformed mean of log(1 + days) with 95% confidence intervals, plotted on a log scale. (B) Distribution of trajectory classes among patients with and without a gap in recorded contact longer than 365 days. (C) Cumulative incidence of cardiovascular hospitalization and (D) of death without such a hospitalization after the landmark, by class (Aalen–Johansen estimates), with Gray’s test.

The two measures largely identified the same patients (**Figure 3B**). Among the 3,777 patients (24.8%) with a contact gap exceeding 365 days, 96% belonged to the Minimal Contact class, and none belonged to the Sustained Contact class. The gap definition did not require a subsequent return; however, inclusion in the primary landmark cohort required a record after day 730. The cumulative incidence of cardiovascular hospitalization varied modestly across contact-pattern classes, ranging from 4.4% in the Minimal Contact class to 5.8% in the Rising Contact class at 730 days (Gray’s test, p = 0.023). The corresponding incidence was 4.2% among patients with a contact gap and 5.1% among those without one (Gray’s test, p = 0.042). In contrast, death without prior cardiovascular hospitalization differed more substantially across classes, increasing from 403 deaths (5.3%) in the Minimal Contact class to 212 (7.9%) in the Rising Contact class and 258 (16.8%) in the Sustained Contact class (Gray’s test, p < 0.001; **Table S7**). These differences in competing mortality are consistent with the greater comorbidity burden and health care utilization observed among patients with more sustained contact (**Figure 3C–D**).

Adjusted estimates are presented in Table 4. A recorded contact gap was not associated with an increased hazard of cardiovascular hospitalization. Furthermore, the duration of the longest interval without recorded contact showed no clear dose–response relationship (overall p = 0.26; Figure S2). Relative to a 90-day interval, the estimated HRs were 0.93 (95% CI, 0.76–1.13) for 180 days, 1.03 (95% CI, 0.81–1.31) for 366 days, and 0.73 (95% CI, 0.51–1.04) for 730 days. All corresponding 95% CIs included the null value of 1. Similarly, a contact gap exceeding 365 days was not associated with a statistically significant increase in the hazard of cardiovascular hospitalization in either the cause-specific model (HR, 0.86; 95% CI, 0.71–1.05) or the Fine–Gray subdistribution hazard model (HR, 0.86; 95% CI, 0.71–1.05). The doubly robust estimate was below 1 (HR, 0.81; 95% CI, 0.66–0.98). Gaps exceeding 180 days (HR, 0.94; 95% CI, 0.79–1.11) or 270 days (HR, 1.03; 95% CI, 0.86–1.22), as well as gaps within the Minimal Contact class (HR, 0.92; 95% CI, 0.72–1.17), were also not associated with higher hazards. Taken together, these findings do not support a consistent association between longer contact gaps and increased cardiovascular hospitalization risk, although the estimates do not establish the absence of an association. The overall association between contact-pattern class and cardiovascular hospitalization was borderline (p = 0.05). Compared with Minimal Contact, Sustained Contact was associated with a higher cause-specific hazard in the covariate-adjusted model (HR, 1.40; 95% CI, 1.06–1.85) and the doubly robust model (HR, 1.43; 95% CI, 1.10–1.85). However, the corresponding Fine–Gray subdistribution HR was less precise and included the null (HR, 1.28; 95% CI, 0.97–1.70). This difference across estimands is relevant given the substantially higher incidence of death without cardiovascular hospitalization in the Sustained Contact class. Rising Contact was associated with a more modest increase in the cause-specific hazard (HR, 1.26; 95% CI, 1.02–1.56), whereas its Fine–Gray estimate was compatible with no association (HR, 1.23; 95% CI, 0.99–1.52). No statistically significant association was observed for Declining Contact relative to Minimal Contact (HR, 1.08; 95% CI, 0.88–1.33).

After weighting, standardized differences were 0.12 or lower across contact-pattern classes, approaching the prespecified balance target of 0.10; the standardized difference for contact-gap status was 0.22. The doubly robust estimates additionally incorporated direct covariate adjustment. Proportional hazards diagnostics provided no strong evidence of violations, with the smallest p value for an exposure term being 0.07. In the day-730-information sample, which excluded post-landmark information, the results were directionally similar but less precise (n = 7,649; 318 cardiovascular hospitalizations). The HR was 0.77 (95% CI, 0.55–1.09) for a contact gap exceeding 365 days and 1.33 (95% CI, 0.92–1.93) for Sustained Contact. The overall association between contact-pattern class and cardiovascular hospitalization was not statistically significant in this restricted sample (p = 0.34; **Table 4**). Neither contact measure materially improved discrimination beyond the baseline covariates (Table 4B). The covariate-only model had a C-statistic of 0.708. Adding contact-gap status increased the C-statistic by 0.0006 (95% CI, −0.0011 to 0.0023; Wald p = 0.14), whereas adding contact-pattern class increased it by 0.0021 (95% CI, −0.0006 to 0.0053; p = 0.05). Including both measures yielded an increase of 0.0023 (95% CI, −0.0006 to 0.0056). Once the other contact measure was included, the incremental contribution of contact-pattern class was not statistically significant (p = 0.10), nor was that of contact-gap status (p = 0.53). Thus, although Sustained Contact and, to a lesser extent, Rising Contact were associated with higher cause-specific hazards of cardiovascular hospitalization, neither measure demonstrated a substantial improvement in discrimination beyond the covariates.

**Table 4.** Events, 730-day cumulative incidence, and adjusted associations for both contact measures in the landmark population (n = 15,200)

| Exposure | n | Events | Deaths without event | 730-day event incidence, % | Cause-specific HR (95% CI) | Fine-Gray sHR (95% CI) | Doubly robust HR (95% CI) |
| --- | --- | --- | --- | --- | --- | --- | --- |
| **Longest interval without contact (restricted cubic spline: overall p = 0.26; Figure S2)** |  |  |  |  |  |  |  |
| 90 days (reference) | — | — | — | — | 1 (reference) | — | — |
| 180 days | — | — | — | — | 0.93 (0.76–1.13) | — | — |
| 366 days | — | — | — | — | 1.03 (0.81–1.31) | — | — |
| 730 days | — | — | — | — | 0.73 (0.51–1.04) | — | — |
| **Gap in recorded contact longer than 365 days (Gray's test: events p = 0.042; deaths p = 0.31)** |  |  |  |  |  |  |  |
| No gap >365 days | 11,423 | 518 | 817 | 5.1 | 1 (reference) | 1 (reference) | 1 (reference) |
| Gap >365 days | 3,777 | 146 | 260 | 4.2 | 0.86 (0.71–1.05) | 0.86 (0.71–1.05) | 0.81 (0.66–0.98) |
| **Trajectory class (overall p = 0.05; Gray's test: events p = 0.023; deaths p < 0.001)** |  |  |  |  |  |  |  |
| Minimal Contact | 7,569 | 303 | 403 | 4.4 | 1 (reference) | 1 (reference) | 1 (reference) |
| Declining Contact | 3,413 | 152 | 204 | 5.0 | 1.08 (0.88–1.33) | 1.08 (0.88–1.33) | 1.09 (0.89–1.32) |
| Rising Contact | 2,679 | 135 | 212 | 5.8 | 1.26 (1.02–1.56) | 1.23 (0.99–1.52) | 1.27 (1.04–1.57) |
| Sustained Contact | 1,539 | 74 | 258 | 5.6 | 1.40 (1.06–1.85) | 1.28 (0.97–1.70) | 1.43 (1.10–1.85) |
| **Day-730-information sample (n = 7,649; 318 events; overall class p = 0.34)<sup>b</sup>** |  |  |  |  |  |  |  |
| Gap >365 days | — | — | — | — | 0.77 (0.55–1.09) | — | — |
| Sustained Contact | — | — | — | — | 1.33 (0.92–1.93) | — | — |

**B. Added information from each measure**
| Model (Uno's C-statistic at 730 days) | C-statistic or change (95% CI) | Pooled Wald p |
| --- | --- | --- |
| Covariates only | 0.708 | — |
| + gap status | +0.0006 (−0.0011 to +0.0023) | 0.14 |
| + trajectory class | +0.0021 (−0.0006 to +0.0053) | 0.05 |
| + gap status and class | +0.0023 (−0.0006 to +0.0056) | — |
| Class, in a model already containing gap status | — | 0.10 |
| Gap status, in a model already containing class | — | 0.53 |

### Sensitivity analyses and outcome components

The findings were generally consistent across sensitivity analyses (**Table S3**). Excluding the complete-case analysis, hazard ratios (HRs) for a contact gap exceeding 365 days ranged from 0.77 to 0.93, whereas estimates for Sustained Contact ranged from 1.28 to 1.43. The overall test for differences across contact-pattern classes yielded p values ranging from 0.02 to 0.34, indicating some variability in the statistical evidence across model specifications. Point estimates were similar across most alternative specifications; the complete-case analysis yielded a lower and imprecise estimate. The HR remained similar when covariates were averaged over the period from 365 days before to 90 days after index (HR, 1.42), when continuous covariates were modeled linearly (HR, 1.39), and when the missing-indicator method was used (HR, 1.40). Delta-adjusted imputation likewise yielded estimates ranging from 1.39 to 1.40. The estimate was also similar when patients with cardiovascular events before index were retained (HR, 1.41; 95% CI, 1.09–1.83; n = 16,446; 798 events) and when posterior class-membership probabilities were used in place of assigned classes (HR, 1.42; 95% CI, 1.05–1.93).

The complete-case analysis was the principal exception, yielding an HR of 1.11 (95% CI, 0.71–1.72) for a contact gap exceeding 365 days and 0.88 (95% CI, 0.53–1.48) for Sustained Contact. This analysis was restricted to the 28% of patients with complete data on all three incompletely observed clinical measures—estimated glomerular filtration rate (eGFR), glycated hemoglobin, and body mass index—and included 184 events. The confidence intervals were wide and included the corresponding primary estimates, limiting the strength of any inference that these results differed from the primary findings. Observed and imputed means were similar for eGFR (76.4 vs. 77.5 mL/min/1.73 m²), body mass index (30.5 vs. 30.2 kg/m²), and glycated hemoglobin (6.69% vs. 6.73%). The fraction of missing information was 0.01 or lower for every estimate, indicating limited loss of statistical information attributable to missingness under the fitted imputation models. Nevertheless, similarity between observed and imputed means and low fractions of missing information do not, by themselves, establish the validity of the missing-data assumptions. With 664 cardiovascular hospitalization events, the study had 80% power to detect an HR of 1.29 (or 0.78) for a contact gap exceeding 365 days and HRs ranging from 1.33 to 1.47 for Declining, Rising, and Sustained Contact relative to Minimal Contact. These calculations suggest that the study was better positioned to detect larger associations than modest ones; therefore, smaller associations cannot be excluded.

In exploratory analyses of individual outcome components (**Table S8; Figure S3**), neither contact gaps nor contact-pattern classes showed evidence of an association with myocardial infarction or stroke. In contrast, the higher hazards observed for Rising and Sustained Contact were concentrated in heart failure hospitalization. Relative to Minimal Contact, Rising Contact was associated with an HR of 1.66 (95% CI, 1.17–2.35), whereas Sustained Contact was associated with an HR of 2.14 (95% CI, 1.38–3.30), based on 283 heart failure hospitalization events. These associations persisted after adjustment for prior heart failure diagnoses and among patients without a documented heart failure diagnosis before the landmark. In the latter subgroup, the HRs were 1.67 (95% CI, 1.01–2.75) for Rising Contact and 2.07 (95% CI, 1.07–4.02) for Sustained Contact. Although these findings suggest that the observed associations were not explained entirely by previously documented heart failure, the exploratory nature of the analyses and the relatively wide confidence intervals warrant cautious interpretation. When diagnoses recorded in any diagnostic position were included, the number of composite outcome events increased more than threefold, and the estimated associations increased with contact intensity. This pattern is consistent with the possibility that patients with more frequent health care encounters have greater opportunities for diagnostic evaluation and coding. Accordingly, the stronger associations observed under this broader outcome definition may partly reflect differential ascertainment rather than differences in underlying disease incidence alone.

## DISCUSSION

In this cohort study of adults with hypertension receiving care within a single academic health system, the absence of a recorded blood pressure measurement within 90 days after an elevated reading was common and could be predicted using information available in the EHRs at the time of the elevated reading. More than one-third of patients had no recorded measurement within this interval. A prediction model relying primarily on health care contact history achieved an AUROC of 0.74 and an AUPRC of 0.57, compared with an outcome prevalence of 0.35. At an outreach capacity of 20%, approximately six in ten patients selected for outreach subsequently had no recorded measurement. In contrast, analyses of cardiovascular hospitalization revealed a different pattern: a prolonged gap in recorded contact was not consistently associated with a higher hazard of cardiovascular hospitalization, whereas Rising and Sustained Contact were associated with modestly higher cause-specific hazards, largely attributable to heart failure hospitalization. When contact measures and covariates were defined before the landmark, incorporating contact patterns or gaps into models based on baseline covariates yielded little improvement in discrimination for cardiovascular hospitalization. Collectively, these findings suggest that EHR-derived contact history may help prioritize blood pressure follow-up, but they do not establish that model-guided outreach improves care or that contact history materially enhances cardiovascular risk prediction.

The most influential predictors of absent recorded follow-up were measures of prior health care utilization, including time since the last visit, the number and spacing of previous encounters, and blood pressure variability. The latter requires multiple measurements and may therefore be unavailable or less informative for patients with limited prior engagement. A parsimonious model using only time since the last visit and the number of prior encounters achieved an AUROC of 0.69, indicating that useful risk stratification may be feasible without a more complex machine learning model.[51] Nevertheless, the full model identified more patients without a subsequent recorded measurement at each outreach-capacity level and yielded positive net benefit across a wider range of decision thresholds. Its discrimination was within the range reported for models predicting missed appointments [52], although the outcomes are not directly equivalent: absence of a recorded blood pressure measurement after an elevated reading reflects an undocumented step in a care process and does not necessarily indicate a missed appointment or failure to obtain care elsewhere. Discrimination improved over longer prediction windows, which may primarily reflect the identification of patients who cease appearing in the health system altogether. The 90-day window may nevertheless be more actionable because it aligns more closely with the recommended interval for reassessment after an elevated reading [5].

Evaluating performance in relation to outreach capacity helps translate discrimination into a potential operational strategy. At outreach capacities of 5% and 30%, approximately six in ten patients selected for hypothetical outreach by the model subsequently had no recorded measurement, compared with approximately one in three patients contacted at random. These results suggest that contact history could help direct limited outreach resources toward patients more likely to lack documented follow-up.[53] However, the outcome used for model development was the absence of a recorded measurement, not the absence of measurement in all care settings, and a positive prediction does not establish that an intervention is necessary or beneficial. Implementation findings from the same program also describe patient registries, team-based care, and improved medication access as components of a broader hypertension-care strategy [54]. The study did not evaluate whether model-guided outreach increased follow-up, improved blood pressure control, or reduced cardiovascular events. Establishing clinical utility will therefore require prospective evaluation, ideally through a randomized trial or a well-designed quasi-experimental study comparing model-guided outreach with usual care.

The subgroup analyses revealed a trade-off that aggregate calibration metrics alone do not capture. Although average predicted and observed risks were concordant across racial and ethnic groups, the model identified 22% of Black patients and 23% of Hispanic patients without a recorded measurement at the evaluated threshold, compared with 37% of White patients. Differences in outcome prevalence can contribute to differences in classification performance even when a model is calibrated within groups [10]. In this study, however, outcome prevalence differed by approximately 4–5 percentage points, whereas detection rates differed by 14–16 percentage points. This disparity suggests that differences in the distribution or recording of predictors, including health care contact history, may also have contributed, consistent with concerns about algorithms that rely on patterns of health care utilization [9]. Applying group-specific thresholds brought detection rates closer together but increased false-positive rates among Black and Hispanic patients. For a relatively low-burden intervention such as a scheduling call, false-positive classifications may carry limited direct costs, although they can still impose burdens on patients and health systems. Using race-specific thresholds nevertheless raises important legal and ethical questions. Alternatives that do not explicitly use race, including expanding outreach capacity or incorporating better measures of access to care, warrant evaluation. We therefore present group-specific thresholds as an illustration of the trade-offs between equity objectives and classification errors, rather than as a recommendation for race-specific deployment. Patients categorized as having another or unknown race or ethnicity—including those whose race was not recorded—had the highest prevalence of absent recorded follow-up and were flagged most frequently. Because missing demographic information may itself be associated with patterns of health care contact, this finding also highlights the potential importance of data completeness when evaluating algorithmic fairness.[55]

For cardiovascular hospitalization, the results suggest that recorded contact history reflects clinical complexity and patterns of health care utilization more directly than it reflects future cardiovascular risk. Contact-pattern classes differed relatively little in age, blood pressure, and laboratory measurements but substantially in comorbidity burden, prior health care utilization, and mortality. Patients with gaps exceeding 365 days were concentrated in the Minimal Contact class, which had the lowest comorbidity burden and fewest prior encounters. For some patients, therefore, a prolonged interval without a recorded encounter may reflect stable disease, lower clinical need, or care received outside the health system rather than inadequate care. The absence of a consistent dose–response relationship between gap duration and cardiovascular hospitalization, together with the limited incremental discrimination provided by contact measures, argues against interpreting a prolonged EHR contact gap as a stand-alone marker of cardiovascular risk. These findings were broadly consistent across alternative estimators and missing-data approaches, and estimates were similar in the sample restricted to information available at the landmark.

These findings differ from previous studies linking patterns of care to clinical outcomes. Greater continuity of care was associated with fewer cardiovascular events in Korean national claims data [15], delayed diagnosis following elevated blood pressure readings was associated with higher cardiovascular risk in a large US health system [6], and missed visits were associated with higher mortality in HIV care, where missed visits were also interpreted as a marker of risk rather than necessarily its cause [56]. Differences in outcome and exposure definitions, population selection, and data capture may help explain the apparent discrepancies. National claims data can capture encounters across multiple providers, making a prolonged gap more informative about overall health care utilization than a gap observed within a single health system. The Korean study also required at least four visits during the first two years, selecting a population with established health care contact, although our landmark design similarly conditioned on evidence of survival through day 730. Delayed diagnosis following elevated blood pressure is more directly related to inaction within a care process than is an EHR-defined gap in contact. In addition, interruption of HIV treatment may have relatively rapid clinical consequences, whereas the cardiovascular consequences of inadequate hypertension follow-up may require a longer period to become apparent. These distinctions underscore the importance of defining contact gaps in relation to the data source, clinical process, and outcome under investigation.

The modestly higher cause-specific hazards observed in the Rising and Sustained Contact classes were largely confined to heart failure hospitalization and persisted after adjustment for prior heart failure diagnoses and among patients without a documented heart failure diagnosis before the landmark. These findings warrant cautious interpretation. The component analyses were exploratory and involved 12 comparisons: contact-gap status and three class contrasts for each of three cardiovascular outcome components. In addition, the association for Sustained Contact was attenuated and less precise in the Fine–Gray model, which estimates the subdistribution hazard in the presence of competing events. One plausible explanation is that increasing contact reflects emerging cardiac disease that prompts additional encounters before a heart failure hospitalization. More frequent encounters may also increase opportunities for heart failure recognition and coding [34]. The persistence of the association among patients without a previously documented diagnosis does not exclude subclinical or previously unrecognized disease. Taken together, these findings are more consistent with contact intensity acting as a marker of underlying or emerging illness and differential diagnostic ascertainment. These associations may reflect underlying illness and diagnostic ascertainment and should not be interpreted causally.

Finally, the study highlights how cohort construction can influence analyses of retention and follow-up using routinely collected EHR data.[57] Studies of retention frequently define eligibility according to whether patients return for subsequent care, thereby conditioning on the behavior under investigation [16, 17]. In our analyses, restricting the prediction sample to patients with subsequent recorded contact reduced the 270-day prevalence of absent recorded blood pressure follow-up to 9.4%, compared with 27.9% among patients excluded by that restriction. This substantial difference illustrates how requiring later contact can select a population with a markedly different follow-up profile and underestimate the frequency of undocumented follow-up in the broader eligible population. Explicitly distinguishing database coverage, evidence of survival, and death ascertainment makes these selection mechanisms more transparent and provides a framework that may be useful in other single-system EHR studies. Defining the cohort at the index reading, however, does not eliminate selection. The cardiovascular analysis remained conditioned on evidence of survival through the day-730 landmark, while the prediction analysis required 90 days of database coverage after the elevated reading. These design choices define the populations to which the respective findings apply and should be considered when interpreting their generalizability.

### Limitations

This study has several limitations. First, care received outside the study health system was not captured; therefore, the absence of a recorded blood pressure measurement does not necessarily indicate absent follow-up care. The single-center MIMIC-IV population may also limit generalizability. Second, the landmark design selected patients with evidence of survival through day 730, and survival ascertainment required subsequent hospital or emergency department contact. Analyses using day-730 information yielded similar but less precise estimates, although selection related to health care utilization remained. Third, uncertainty in contact-pattern classification, missing clinical measurements, limited socioeconomic data, and unavailable cause of death may have affected estimates. With 664 cardiovascular hospitalization events, moderate associations cannot be excluded. Finally, the prediction model underwent internal validation using a random split and nested cross-validation, without temporal or external validation, and incomplete race and ethnicity data limited the assessment of algorithmic fairness.

Future studies should validate the model temporally and across health systems, evaluate model-guided outreach in randomized or quasi-experimental studies, and investigate approaches to reduce disparities in detection without relying on race-specific thresholds.[58, 59]

## CONCLUSIONS

Among adults with hypertension, routinely collected EHR data provided moderate discrimination for the absence of a recorded blood pressure measurement within 90 days after an elevated reading. At a simulated outreach capacity of 20%, approximately 60% of selected patients had no recorded follow-up measurement. However, detection varied across racial, ethnic, and age groups, and absent documentation does not establish that care was missed. Recorded contact patterns added little discrimination for subsequent cardiovascular hospitalization. These findings support further evaluation of contact history for prioritizing blood pressure reassessment. External validation and prospective evaluation are needed to determine whether model-guided outreach improves follow-up and blood pressure control while addressing disparities in detection.

## Data Availability

All data produced are available online at https://mimic.physionet.org/.

## DECLARATIONS

### Ethics approval and consent to participate

Not applicable.

### Consent for publication

This study used data from the MIMIC-IV database, a publicly available, de-identified database. As the study involved secondary analysis of existing de-identified data and did not involve direct interaction with human participants or access to identifiable private information, informed consent and Institutional Review Board (IRB) approval were waived.

### Availability of data and materials

All data are available at Medical Information Mart for Intensive Care: https://mimic.physionet.org/. The code is available at: https://github.com/RonveerSandhu/htn-bp-followup-mimic.git

### Authors’ contributions

JY developed the study. RS and JY contributed to the data analyses. RS, SA, MH, and JY contributed to the interpretation of the results and the writing of the manuscript. All authors approved the final version of the manuscript.

## Acknowledgements

None.

## SUPPLEMENT

### Supplementary Methods

#### Cohort definition

MIMIC-IV shifts patient dates and provides calendar time only in three-year groups; the latest hospital and emergency records were from 2020. Database coverage was defined using the latest date guaranteed to be available within each patient’s calendar group. Initial eligibility was based on the index criteria and estimated database coverage. Additional survival and record-availability requirements were applied separately to the prediction and landmark analyses, as detailed in **Table S9**. Overall, 7.3% had an admission beginning on the index date, and 99% of recorded deaths occurred within 355 days of the last hospital or emergency contact.

#### Prediction targets and model development

Alternative prediction targets were defined as the absence of a blood pressure measurement within 30, 180, 270, or 365 days, each with target-specific coverage and survival requirements. Competing-risk cumulative incidence of measurement and death before measurement was assessed across predicted-risk fifths. Predictors were restricted to information available at prediction. Training and test assignments were aligned across the 90- and 270-day targets. Logistic regression and tree-based models were tuned using five-fold cross-validation and AUPRC. Imputation and standardization were fitted within training folds; tree models handled missing values directly. Nested cross-validation used five outer folds and three-fold inner tuning, with isotonic recalibration that excluded each patient’s own outcome. Subgroup confidence intervals were estimated using 500 bootstrap samples.

#### Trajectory models

Trajectory models used quadratic time trends, class-specific means, Gaussian errors, and no random effects. One-to six-class solutions were fitted using 20 random starts. Selection considered the Bayesian information criterion, class size, posterior classification probabilities, and classification odds. The four-class solution was the smallest that distinguished rising from declining contact while meeting prespecified classification criteria. Alternative definitions and expanded cohorts were evaluated in sensitivity analyses without refitting the original model.

#### Covariates and missing data

Index characteristics, comorbidities, blood pressure, prior encounters, medication counts, and laboratory measures were derived from records available within specified observation windows. Continuous covariates were modeled using restricted cubic splines or linear terms, as appropriate. Multiple imputation incorporated covariates, contact measures, event indicators, and Nelson–Aalen cumulative hazards. Imputed values were restricted to observed-value percentiles, and delta-adjusted imputation and alternative covariate specifications were evaluated in sensitivity analyses.

#### Gap measure and association analyses

The longest interval without recorded contact was modeled using restricted cubic splines, with knots at the 5th, 35th, 65th, and 95th percentiles. A gap exceeding 365 days was evaluated as a prolonged interval without recorded contact; it does not establish that scheduled follow-up visits were missed. Covariate balance was assessed using standardized mean differences, with 0.1 as the target. Censoring weights were estimated using pooled logistic regression across 91-day intervals and combined across intervals. Weighted models used robust standard errors. Uno’s C-statistics treated death as censoring and were averaged across imputations; confidence intervals used 1,000 bootstrap samples. Discrimination estimates were apparent and not corrected for optimism.

#### Multimorbidity

Comorbidity phenotypes were derived from a co-occurrence network of Elixhauser categories, excluding hypertension, in a cohort requiring post-day-730 records. Categories present in at least 1% of patients were connected when their phi coefficient exceeded 0.10, and Louvain community detection identified three multimorbidity groups: cardio-renal-metabolic, neuro-psychiatric-hepatic, and malignancy. Patients were assigned to the group containing the most of their comorbidities; no group-associated comorbidity. Phenotype distributions were compared using the chi-square test and Cramér’s V, and effect modification was assessed using phenotype-by-contact-class interactions in the cause-specific model.

#### Software

Analyses used Python 3.13 and R 4.6, with packages for data processing, prediction, survival analysis, competing risks, trajectory modeling, explainability, and comorbidity scoring. MIMIC-IV is available to credentialed researchers through PhysioNet. Analysis code is available in the repository specified under Availability of Data and Materials.

### Supplementary Results

**Table S1.** Diagnosis codes and antihypertensive drugs.

A. ICD code prefixes (MIMIC-IV format, without decimal points)
| Definition | ICD-9 | ICD-10 |
| --- | --- | --- |
| Hypertension (inclusion) | 401, 402, 403, 404 | I10, I11, I12, I13 |
| Surgically correctable secondary cause (exclusion) | 4401, 2270, 1940, 2550, 25510–25514, 7471 | I701, E275, C74, D35, E260, E24, Q251 |
| Myocardial infarction | 410 | I21, I22 |
| Stroke | 430, 431, 432, 433, 434, 436 | I60, I61, I62, I63 |
| Heart failure | 428 | I50 |

B. Antihypertensive drugs (matched by name in prescription orders)
| Class | Drugs |
| --- | --- |
| ACE inhibitors | lisinopril, enalapril, ramipril, captopril, benazepril, quinapril, fosinopril |
| Angiotensin receptor blockers | losartan, valsartan, olmesartan, irbesartan, candesartan, telmisartan |
| Beta-blockers | metoprolol, atenolol, carvedilol, labetalol, propranolol, bisoprolol, nebivolol |
| Calcium channel blockers | amlodipine, nifedipine, diltiazem, verapamil, nicardipine, felodipine |
| Thiazide-type diuretics | hydrochlorothiazide, chlorthalidone, indapamide, metolazone |
| Loop diuretics | furosemide, torsemide, bumetanide |
| Potassium-sparing diuretics | spironolactone, eplerenone, triamterene, amiloride |
| Central agents and direct vasodilators | clonidine, methyldopa, hydralazine, minoxidil |
| Alpha-blockers | doxazosin, terazosin, prazosin |

Codes are matched by prefix. Outcomes use the principal diagnosis of an admission (any position in sensitivity analyses). ICD-9 code 412 (old myocardial infarction) is not included.

**Table S2.** Group-based trajectory models: fit by number of classes and classification quality of the selected model.

One- to six-class solutions (fitting sample: landmark population, n = 15,200; all converged)
| Classes | Parameters | Log-likelihood | AIC | BIC | Entropy | Smallest class, % | Lowest class-average posterior | Maximum posterior <0.70, % |
| --- | --- | --- | --- | --- | --- | --- | --- | --- |
| 1 | 4 | -111,037.1 | 222,082.1 | 222,112.6 | — | — | — | — |
| 2 | 8 | -97,378.0 | 194,772.0 | 194,833.1 | 0.85 | 30.1 | 0.93 | 5.2 |
| 3 | 12 | -93,783.1 | 187,590.2 | 187,681.8 | 0.81 | 9.8 | 0.89 | 11.1 |
| 4 (selected) | 16 | -93,104.8 | 186,241.7 | 186,363.7 | 0.74 | 10.1 | 0.74 | 23.0 |
| 5 | 20 | -92,026.4 | 184,092.8 | 184,245.4 | 0.75 | 2.5 | 0.73 | 26.0 |
| 6 | 24 | -91,614.6 | 183,277.2 | 183,460.3 | 0.77 | 2.9 | 0.71 | 23.7 |

**B. Four-class solution by class**
| Class | n (%) | Mean unique contact days per quarter, quarters 1–7 | Change, quarter 7 minus quarter 1 | Average posterior probability | Odds of correct classification |
| --- | --- | --- | --- | --- | --- |
| Minimal Contact | 7,569 (49.8) | 0.56 | -0.01 | 0.92 | 11.7 |
| Declining Contact | 3,413 (22.5) | 1.99 | -1.86 | 0.74 | 9.9 |
| Rising Contact | 2,679 (17.6) | 2.24 | +2.34 | 0.75 | 13.9 |
| Sustained Contact | 1,539 (10.1) | 5.72 | -1.27 | 0.91 | 85.3 |

**Table S3.** Sensitivity analyses: hazard ratios (95% CI) for cardiovascular hospitalization.

| Analysis | n | Events | Gap >365 days | Sustained Contact<sup>c</sup> | Overall class test, p |
| --- | --- | --- | --- | --- | --- |
| **Estimators (landmark population, 30 imputations)** |  |  |  |  |  |
| Primary: cause-specific Cox | 15,200 | 664 | 0.86 (0.71–1.05) | 1.40 (1.06–1.85) | 0.05 |
| Fine–Gray | 15,200 | 664 | 0.86 (0.71–1.05) | 1.28 (0.97–1.70) | 0.16 |
| Doubly robust (weights + covariates) | 15,200 | 664 | 0.81 (0.66–0.98) | 1.43 (1.10–1.85) | 0.02 |
| Class and gap in the same model | 15,200 | 664 | 0.93 (0.75–1.16) | 1.36 (1.01–1.82) | 0.10 |
| **Gap definition** |  |  |  |  |  |
| Longest interval, restricted cubic spline (overall p) | 15,200 | 664 | p = 0.26 | — | — |
| Gap >180 days | 15,200 | 664 | 0.94 (0.79–1.11)<sup>a</sup> | — | — |
| Gap >270 days | 15,200 | 664 | 1.03 (0.86–1.22)<sup>a</sup> | — | — |
| Gap >365 days within Minimal Contact | 7,569 | 303 | 0.92 (0.72–1.17) | — | — |
| **Class definition** |  |  |  |  |  |
| Posterior probabilities in place of class assignment | 15,200 | 664 | — | 1.42 (1.05–1.93) | 0.11 |
| Five-class solution (Sustained Moderate Contact)<sup>c</sup> | 15,200 | 664 | — | 1.41 (1.11–1.79) | 0.05 |
| **Covariates and missing data** |  |  |  |  |  |
| Covariates averaged from –365 to +90 days, missing indicators | 15,200 | 664 | 0.84 (0.69–1.03) | 1.42 (1.07–1.88) | 0.04 |
| Linear covariates (no splines) | 15,200 | 664 | 0.88 (0.72–1.07) | 1.39 (1.05–1.83) | 0.05 |
| Missing-indicator method | 15,200 | 664 | 0.85 (0.69–1.03) | 1.40 (1.06–1.86) | 0.04 |
| Complete cases | 4,244 | 184 | 1.11 (0.71–1.72) | 0.88 (0.53–1.48) | 0.96 |
| Missing not at random, 5 delta scenarios (range) | 15,200 | 664 | 0.86 | 1.39–1.40 | 0.04–0.05 |
| **Population and follow-up** |  |  |  |  |  |
| Survival confirmed with day-730 information only<sup>b</sup> | 7,649 | 318 | 0.77 (0.55–1.09) | 1.33 (0.92–1.93) | 0.34 |
| Events before index kept<sup>b</sup> | 16,446 | 798 | 0.83 (0.70–1.00) | 1.41 (1.09–1.83) | 0.02 |
| Censoring weights, truncated at 1st/99th percentiles | 15,200 | 664 | 0.86 (0.71–1.05) | 1.39 (1.04–1.84) | — |
| Censoring weights, truncated at 5th/95th percentiles | 15,200 | 664 | 0.86 (0.71–1.05) | 1.39 (1.05–1.84) | — |
| Censoring at the end of database coverage only | 15,200 | 664 | 0.88 (0.72–1.07) | 1.33 (1.01–1.76) | 0.13 |

**Table S4.** Characteristics of the prediction sample by data set.

| Characteristic | Training | Validation | Test |
| --- | --- | --- | --- |
| Patients, n | 18,564 | 6,189 | 6,189 |
| No recorded BP within 90 days, n (%) | 6,526 (35.2) | 2,188 (35.4) | 2,185 (35.3) |
| Age, years, mean (SD) | 66.6 (13.6) | 66.6 (13.5) | 66.8 (13.7) |
| Female, n (%) | 9,624 (51.8) | 3,220 (52.0) | 3,188 (51.5) |
| **Race and ethnicity, n (%)** |  |  |  |
| White | 12,197 (65.7) | 4,077 (65.9) | 4,070 (65.8) |
| Black | 3,491 (18.8) | 1,174 (19.0) | 1,127 (18.2) |
| Hispanic | 1,021 (5.5) | 335 (5.4) | 354 (5.7) |
| Asian | 612 (3.3) | 184 (3.0) | 206 (3.3) |
| Other or unknown | 1,243 (6.7) | 419 (6.8) | 432 (7.0) |
| **Insurance, n (%)** |  |  |  |
| Medicare | 8,483 (45.7) | 2,861 (46.2) | 2,881 (46.6) |
| Medicaid | 1,162 (6.3) | 384 (6.2) | 380 (6.1) |
| Other | 8,919 (48.0) | 2,944 (47.6) | 2,928 (47.3) |
| **Clinical** |  |  |  |
| Systolic BP, earlier readings, mmHg, mean (SD) | 132.2 (12.9) | 132.4 (12.9) | 132.2 (13.3) |
| eGFR, mL/min/1.73 m<sup>2</sup>, median (IQR) | 77 (56–94) | 77 (55–94) | 76 (56–93) |
| HbA1c, %, median (IQR) | 6.1 (5.7–7.1) | 6.1 (5.7–7.0) | 6.1 (5.7–7.2) |
| BMI, kg/m<sup>2</sup>, mean (SD) | 30.3 (7.2) | 30.1 (7.0) | 30.1 (7.1) |
| Elixhauser score, median (IQR) | 5 (0–11) | 5 (0–12) | 5 (0–11) |
| Antihypertensive drugs, median (IQR) | 2 (0–3) | 2 (0–3) | 2 (0–3) |
| **Contact history before prediction** |  |  |  |
| Prior encounters, median (IQR) | 6 (3–13) | 6 (3–13) | 6 (3–13) |
| Any prior emergency visit, n (%) | 2,576 (13.9) | 846 (13.7) | 855 (13.8) |
| Days since last visit, median (IQR) | 30 (13–83) | 29 (13–82) | 30 (13–78) |
| **Missing values, n (%)** |  |  |  |
| eGFR | 1,046 (5.6) | 331 (5.3) | 334 (5.4) |
| HbA1c | 10,886 (58.6) | 3,693 (59.7) | 3,643 (58.9) |
| BMI | 3,521 (19.0) | 1,215 (19.6) | 1,206 (19.5) |
| Days since last visit | 1,889 (10.2) | 624 (10.1) | 687 (11.1) |
| Systolic BP variability (fewer than two readings) | 5,763 (31.0) | 1,946 (31.4) | 1,959 (31.7) |

**Table S5.** Prediction of no recorded blood pressure measurement: alternative targets and death as a competing event.

Alternative targets (five-fold out-of-fold predictions)
| Target | n | Outcome, n (%) | AUROC (95% CI) | AUPRC (95% CI) | AUROC of the 90-day scores |
| --- | --- | --- | --- | --- | --- |
| No BP within 30 days | 31,278 | 18,608 (59.5) | 0.737 (0.731–0.742) | 0.776 (0.768–0.783) | 0.725 |
| No BP within 90 days (primary) | 30,942 | 10,899 (35.2) | 0.747 (0.741–0.752) | 0.579 (0.569–0.589) | — |
| No BP within 180 days | 30,081 | 6,878 (22.9) | 0.770 (0.764–0.776) | 0.468 (0.455–0.481) | 0.760 |
| No BP within 270 days | 29,127 | 5,172 (17.8) | 0.786 (0.779–0.792) | 0.417 (0.403–0.431) | 0.759 |
| No BP within 365 days | 27,792 | 4,378 (15.8) | 0.791 (0.785–0.798) | 0.393 (0.379–0.408) | 0.759 |
| No BP within 90 days, deaths before BP counted | 31,016 | 10,973 (35.4) | 0.745 (0.740–0.750) | 0.578 (0.569–0.589) | 0.746 |

**B. Cumulative incidence within 90 days of prediction by fifth of predicted risk (n = 31,390)**
| Risk fifth | n | BP reading, % | Death before any BP reading, % | Neither, % |
| --- | --- | --- | --- | --- |
| 1 (lowest) | 6,278 | 91.5 | 0.4 | 8.1 |
| 2 | 6,278 | 77.8 | 0.3 | 21.9 |
| 3 | 6,278 | 65.6 | 0.2 | 34.2 |
| 4 | 6,278 | 50.8 | 0.1 | 49.1 |
| 5 (highest) | 6,278 | 38.0 | 0.2 | 61.8 |

**Table S6.**
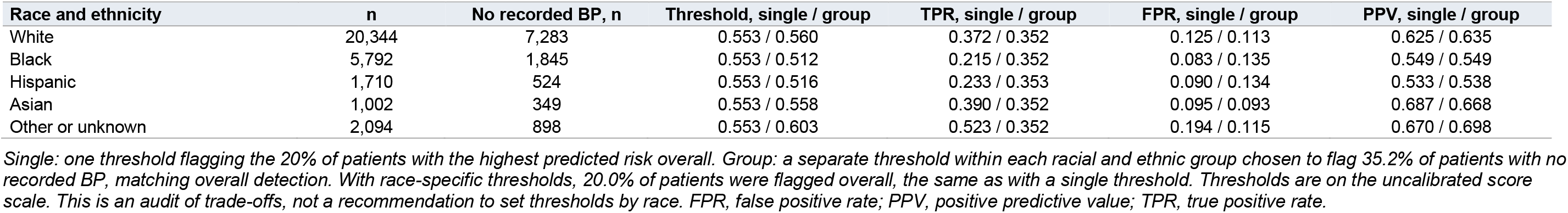
Detection with a single threshold versus race-specific thresholds (out-of-fold predictions, n = 30,942)

**Table S7.** Baseline characteristics of the landmark population, overall and by trajectory class.

| Characteristic | Overall | Minimal Contact | Declining Contact | Rising Contact | Sustained Contact | Max SMD |
| --- | --- | --- | --- | --- | --- | --- |
| Patients, n | 15,200 | 7,569 | 3,413 | 2,679 | 1,539 |  |
| Age, years, mean (SD) | 64.8 (14.0) | 64.6 (14.2) | 65.3 (14.0) | 65.6 (13.9) | 63.3 (13.2) | 0.17 |
| Female, n (%) | 8,282 (54.5) | 3,888 (51.4) | 1,967 (57.6) | 1,564 (58.4) | 863 (56.1) | 0.14 |
| **Race and ethnicity, n (%)** |  |  |  |  |  |  |
| White | 9,679 (63.7) | 5,024 (66.4) | 2,065 (60.5) | 1,608 (60.0) | 982 (63.8) | 0.13 |
| Black | 3,338 (22.0) | 1,495 (19.8) | 847 (24.8) | 660 (24.6) | 336 (21.8) | 0.12 |
| Hispanic | 1,058 (7.0) | 444 (5.9) | 261 (7.6) | 233 (8.7) | 120 (7.8) | 0.11 |
| Asian | 466 (3.1) | 236 (3.1) | 99 (2.9) | 78 (2.9) | 53 (3.4) | 0.03 |
| Other or unknown | 659 (4.3) | 370 (4.9) | 141 (4.1) | 100 (3.7) | 48 (3.1) | 0.09 |
| **Insurance, n (%)** |  |  |  |  |  |  |
| Medicare | 7,138 (47.0) | 3,421 (45.2) | 1,636 (47.9) | 1,394 (52.0) | 687 (44.6) | 0.15 |
| Medicaid | 1,071 (7.0) | 472 (6.2) | 273 (8.0) | 199 (7.4) | 127 (8.3) | 0.08 |
| Other | 6,991 (46.0) | 3,676 (48.6) | 1,504 (44.1) | 1,086 (40.5) | 725 (47.1) | 0.16 |
| **Index measurements** |  |  |  |  |  |  |
| Systolic BP, mmHg, mean (SD) | 131.8 (18.3) | 132.1 (18.2) | 131.8 (18.7) | 131.7 (17.9) | 130.7 (19.0) | 0.07 |
| Diastolic BP, mmHg, mean (SD) | 75.4 (12.2) | 75.9 (12.2) | 75.2 (12.1) | 74.8 (12.0) | 74.7 (12.9) | 0.09 |
| eGFR, mL/min/1.73 m<sup>2</sup>, median (IQR) | 79 (60–96) | 81 (62–97) | 79 (58–95) | 76 (57–95) | 81 (60–98) | 0.13 |
| HbA1c, %, median (IQR) | 6.2 (5.7–7.1) | 6.1 (5.7–7.1) | 6.2 (5.7–7.2) | 6.3 (5.8–7.1) | 6.2 (5.7–7.0) | 0.11 |
| BMI, kg/m<sup>2</sup>, mean (SD) | 30.5 (7.4) | 30.4 (7.3) | 30.5 (7.6) | 30.6 (7.2) | 30.5 (7.5) | 0.02 |
| Elixhauser score, median (IQR) | 4 (0–10) | 4 (0–9) | 5 (0–10) | 4 (0–10) | 7 (2–15) | 0.49 |
| Antihypertensive drugs, median (IQR) | 1 (0–3) | 1 (0–3) | 1 (0–3) | 1 (0–3) | 2 (1–3) | 0.16 |
| **Prior use** |  |  |  |  |  |  |
| Any emergency visit, n (%) | 1,949 (12.8) | 808 (10.7) | 515 (15.1) | 318 (11.9) | 308 (20.0) | 0.26 |
| Two or more inpatient stays, n (%) | 4,925 (32.4) | 2,444 (32.3) | 1,079 (31.6) | 842 (31.4) | 560 (36.4) | 0.10 |
| All encounters, median (IQR) | 4 (2–8) | 3 (2–5) | 6 (3–11) | 5 (3–10) | 9 (4–22) | 0.71 |
| Admission beginning on index date, n (%) | 1,114 (7.3) | 356 (4.7) | 313 (9.2) | 207 (7.7) | 238 (15.5) | 0.36 |
| **Missing values, n (%)** |  |  |  |  |  |  |
| eGFR | 844 (5.6) | 639 (8.4) | 97 (2.8) | 93 (3.5) | 15 (1.0) | 0.36 |
| HbA1c | 9,258 (60.9) | 5,003 (66.1) | 1,909 (55.9) | 1,478 (55.2) | 868 (56.4) | 0.23 |
| BMI | 4,691 (30.9) | 2,939 (38.8) | 812 (23.8) | 737 (27.5) | 203 (13.2) | 0.61 |
| **Contact, days 0–730** |  |  |  |  |  |  |
| Longest interval without contact, days, median (IQR) | 206 (134–365) | 359 (241–526) | 149 (119–187) | 162 (123–210) | 79 (58–98) | 2.37 |
| Gap >365 days, n (%) | 3,777 (24.8) | 3,610 (47.7) | 64 (1.9) | 103 (3.8) | 0 (0.0) | 1.35 |
| **Outcomes within 730 days after the landmark** |  |  |  |  |  |  |
| Cardiovascular hospitalization, n (%) | 664 (4.4) | 303 (4.0) | 152 (4.5) | 135 (5.0) | 74 (4.8) | 0.05 |
| Death without cardiovascular hospitalization, n (%) | 1,077 (7.1) | 403 (5.3) | 204 (6.0) | 212 (7.9) | 258 (16.8) | 0.37 |
| Follow-up, days, median (IQR) | 730 (552–730) | 730 (621–730) | 730 (586–730) | 730 (422–730) | 730 (366–730) |  |

**Table S8.** Outcome components and heart failure sensitivity analyses: cause-specific hazard ratios (95% CI)

Components by diagnosis position
| Outcome | Events | Gap >365 days | Declining Contact | Rising Contact | Sustained Contact |
| --- | --- | --- | --- | --- | --- |
| **Principal diagnosis (primary definition)** |  |  |  |  |  |
| Composite | 664 | 0.86 (0.71–1.05) | 1.08 (0.88–1.33) | 1.26 (1.02–1.56) | 1.40 (1.06–1.85) |
| Myocardial infarction | 197 | 0.96 (0.68–1.35) | 0.83 (0.56–1.22) | 1.10 (0.75–1.61) | 0.69 (0.36–1.30) |
| Stroke | 209 | 0.95 (0.67–1.35) | 1.07 (0.75–1.53) | 0.99 (0.67–1.47) | 1.24 (0.74–2.07) |
| Heart failure | 283 | 0.74 (0.53–1.03) | 1.34 (0.97–1.87) | 1.66 (1.17–2.35) | 2.14 (1.38–3.30) |
| **Any diagnosis position** |  |  |  |  |  |
| Composite | 2,253 | 1.01 (0.91–1.12) | 1.08 (0.96–1.21) | 1.38 (1.23–1.55) | 1.69 (1.46–1.95) |
| Myocardial infarction | 387 | 1.06 (0.83–1.35) | 0.93 (0.71–1.23) | 1.31 (1.00–1.71) | 0.99 (0.66–1.47) |
| Stroke | 387 | 0.94 (0.73–1.21) | 1.17 (0.90–1.53) | 1.05 (0.78–1.41) | 1.58 (1.11–2.26) |
| Heart failure | 1,837 | 1.04 (0.93–1.17) | 1.12 (0.98–1.27) | 1.44 (1.27–1.64) | 1.74 (1.48–2.04) |

**B. Heart failure hospitalization (principal diagnosis): adjustment for prior heart failure**
| Model | n | Events | Rising Contact | Sustained Contact |
| --- | --- | --- | --- | --- |
| Primary model | 15,200 | 283 | 1.66 (1.17–2.35) | 2.14 (1.38–3.30) |
| + baseline heart failure and renal disease | 15,200 | 283 | 1.65 (1.19–2.28) | 2.41 (1.62–3.60) |
| + any heart failure diagnosis before the landmark | 15,200 | 283 | 1.48 (1.07–2.04) | 1.97 (1.32–2.93) |
| No heart failure at baseline or before the landmark | 13,125 | 143 | 1.67 (1.01–2.75) | 2.07 (1.07–4.02) |

**Table S9.**
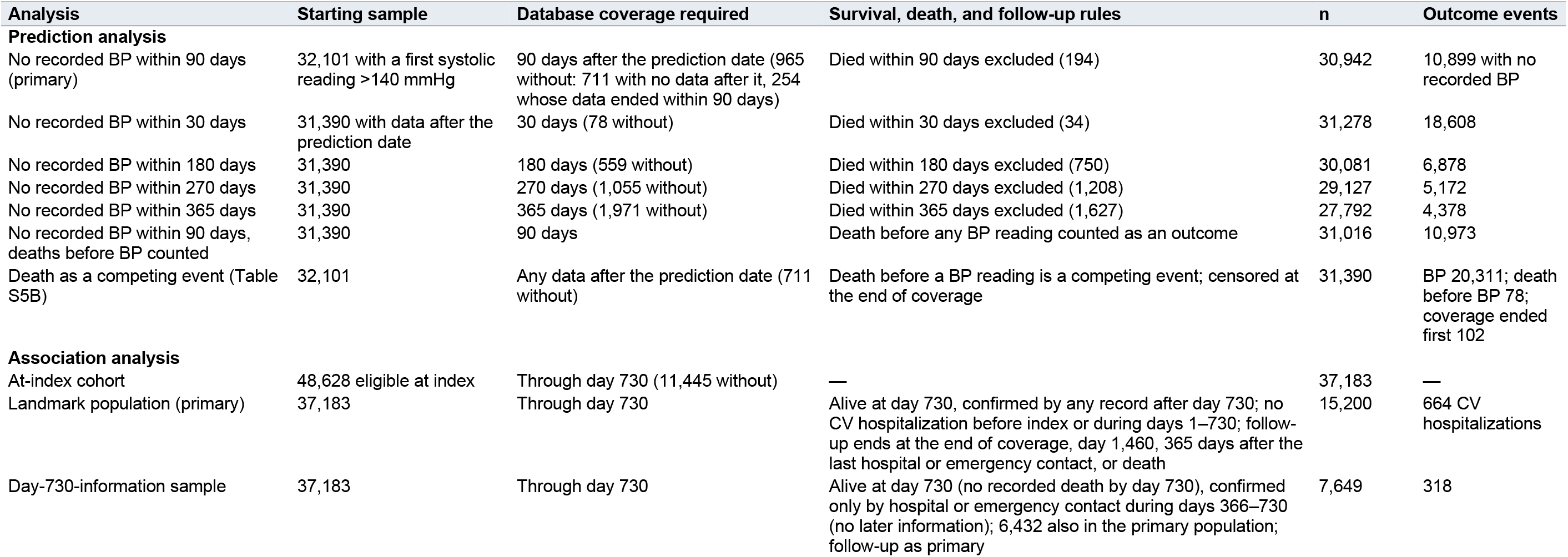
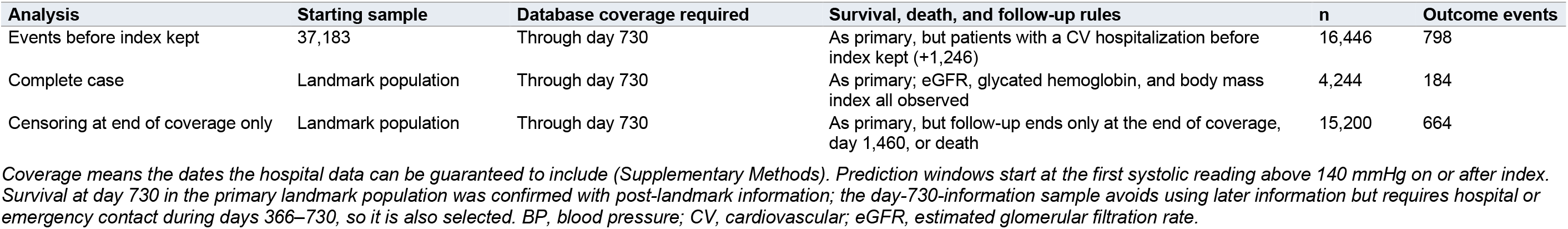
Analysis samples and denominators, with coverage, survival, and death rules.

| Analysis | Starting sample | Database coverage required | Survival, death, and follow-up rules | n | Outcome events |
| --- | --- | --- | --- | --- | --- |
| **Prediction analysis** |  |  |  |  |  |
| No recorded BP within 90 days (primary) | 32,101 with a first systolic reading >140 mmHg | 90 days after the prediction date (965 without: 711 with no data after it, 254 whose data ended within 90 days) | Died within 90 days excluded (194) | 30,942 | 10,899 with no recorded BP |
| No recorded BP within 30 days | 31,390 with data after the prediction date | 30 days (78 without) | Died within 30 days excluded (34) | 31,278 | 18,608 |
| No recorded BP within 180 days | 31,390 | 180 days (559 without) | Died within 180 days excluded (750) | 30,081 | 6,878 |
| No recorded BP within 270 days | 31,390 | 270 days (1,055 without) | Died within 270 days excluded (1,208) | 29,127 | 5,172 |
| No recorded BP within 365 days | 31,390 | 365 days (1,971 without) | Died within 365 days excluded (1,627) | 27,792 | 4,378 |
| No recorded BP within 90 days, deaths before BP counted | 31,390 | 90 days | Death before any BP reading counted as an outcome | 31,016 | 10,973 |
| Death as a competing event (Table S5B) | 32,101 | Any data after the prediction date (711 without) | Death before a BP reading is a competing event; censored at the end of coverage | 31,390 | BP 20,311; death before BP 78; coverage ended first 102 |
| **Association analysis** |  |  |  |  |  |
| At-index cohort | 48,628 eligible at index | Through day 730 (11,445 without) | — | 37,183 | — |
| Landmark population (primary) | 37,183 | Through day 730 | Alive at day 730, confirmed by any record after day 730; no CV hospitalization before index or during days 1–730; follow-up ends at the end of coverage, day 1,460, 365 days after the last hospital or emergency contact, or death | 15,200 | 664 CV hospitalizations |
| Day-730-information sample | 37,183 | Through day 730 | Alive at day 730 (no recorded death by day 730), confirmed only by hospital or emergency contact during days 366–730 (no later information); 6,432 also in the primary population; follow-up as primary | 7,649 | 318 |
| Events before index kept | 37,183 | Through day 730 | As primary, but patients with a CV hospitalization before index kept (+1,246) | 16,446 | 798 |
| Complete case | Landmark population | Through day 730 | As primary; eGFR, glycated hemoglobin, and body mass index all observed | 4,244 | 184 |
| Censoring at end of coverage only | Landmark population | Through day 730 | As primary, but follow-up ends only at the end of coverage, day 1,460, or death | 15,200 | 664 |

*Coverage means the dates the hospital data can be guaranteed to include (Supplementary Methods). Prediction windows start at the first systolic reading above 140 mmHg on or after index. Survival at day 730 in the primary landmark population was confirmed with post-landmark information; the day-730-information sample avoids using later information but requires hospital or emergency contact during days 366–730, so it is also selected. BP, blood pressure; CV, cardiovascular; eGFR, estimated glomerular filtration rate.*

**Figure S1.**
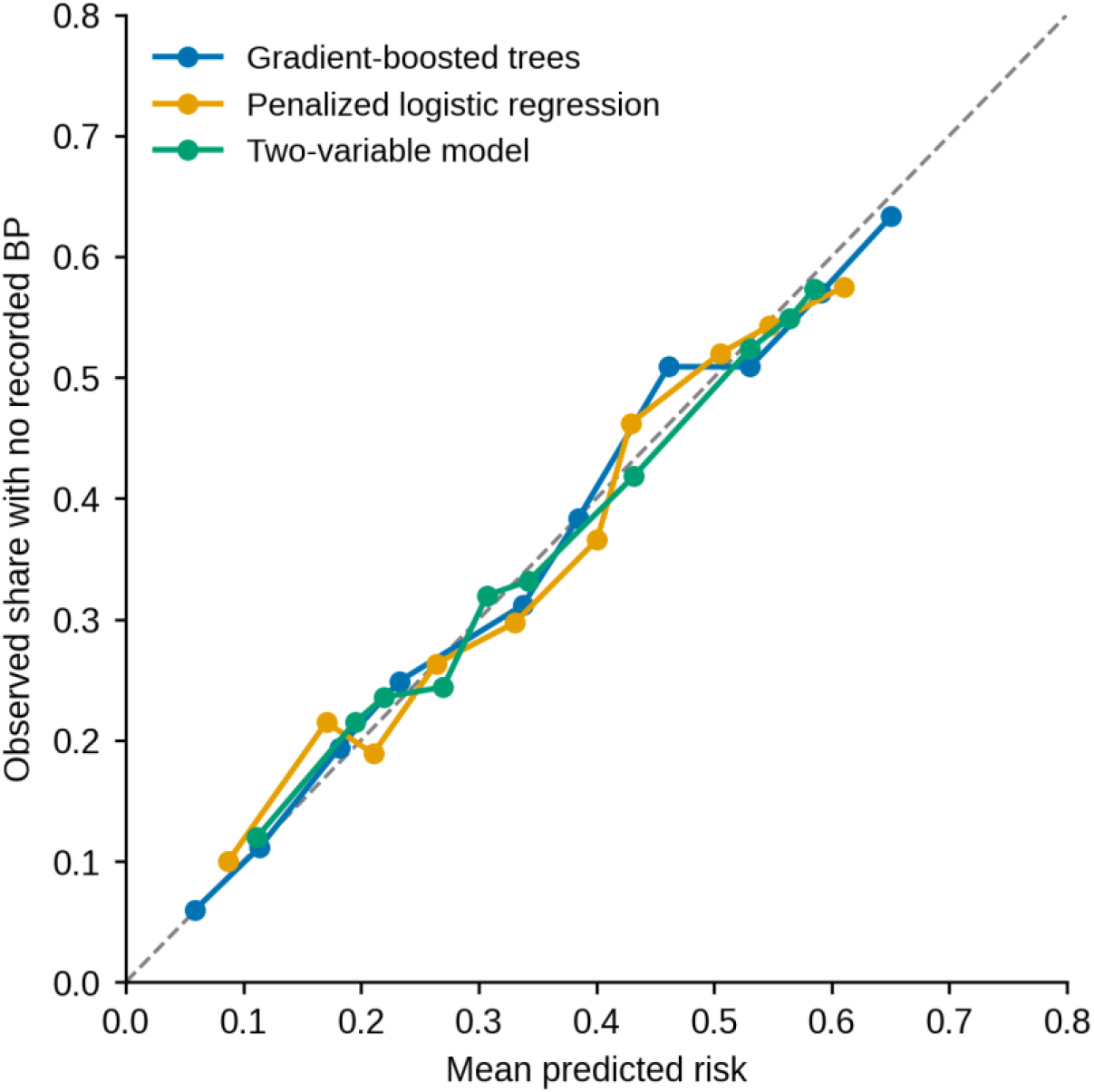
Calibration of the models predicting no recorded blood pressure measurement within 90 days in the test set (n = 6,189). Observed proportions of patients without a recorded blood pressure measurement are plotted against mean predicted risks within deciles of recalibrated predictions.

**Figure S2.**
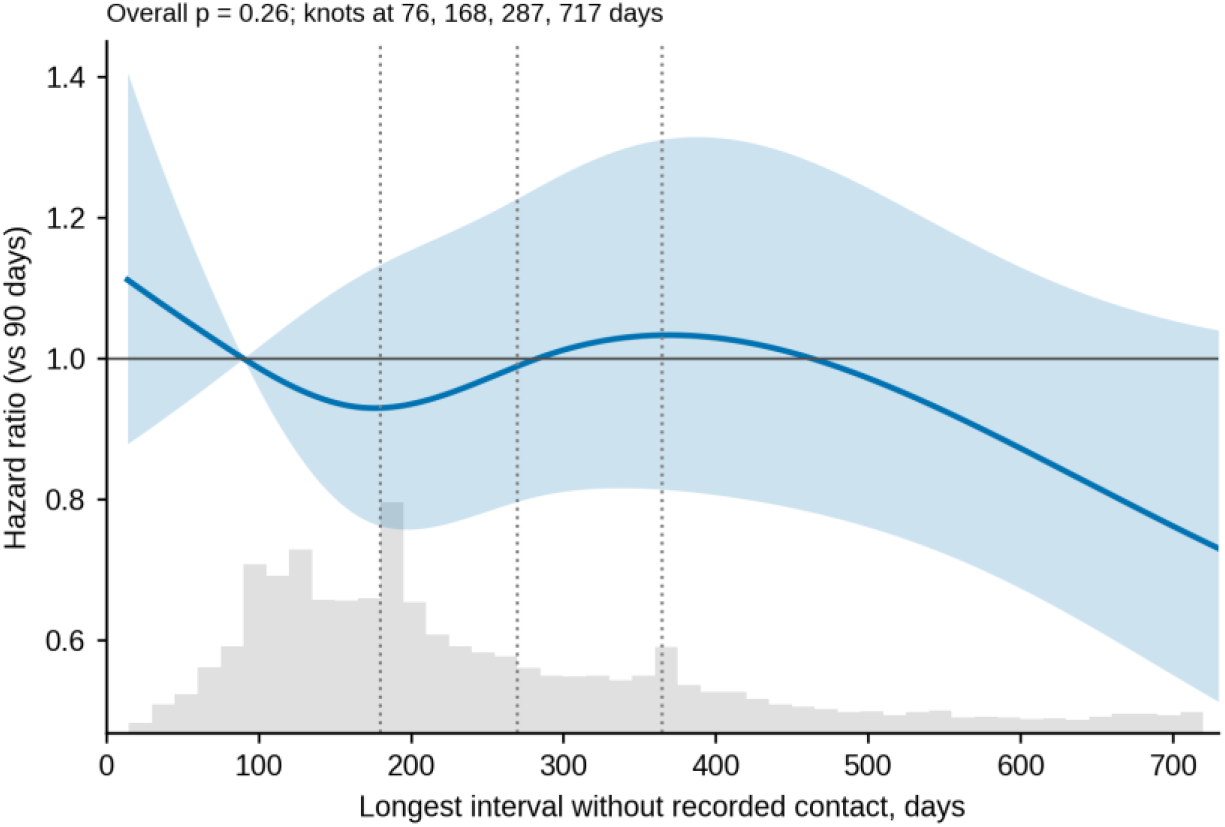
Longest interval without recorded contact and cardiovascular hospitalization. Hazard ratio relative to a 90-day interval from a restricted cubic spline (knots at 76, 168, 287, and 717 days) in a cause-specific Cox model adjusted for the covariates in Table S7, with estimates pooled over 30 imputations (overall p = 0.26). The shaded band is the 95% confidence interval, dotted lines mark 180, 270, and 365 days, and the histogram shows the distribution of intervals in the landmark population (n = 15,200).

**Figure S3.**
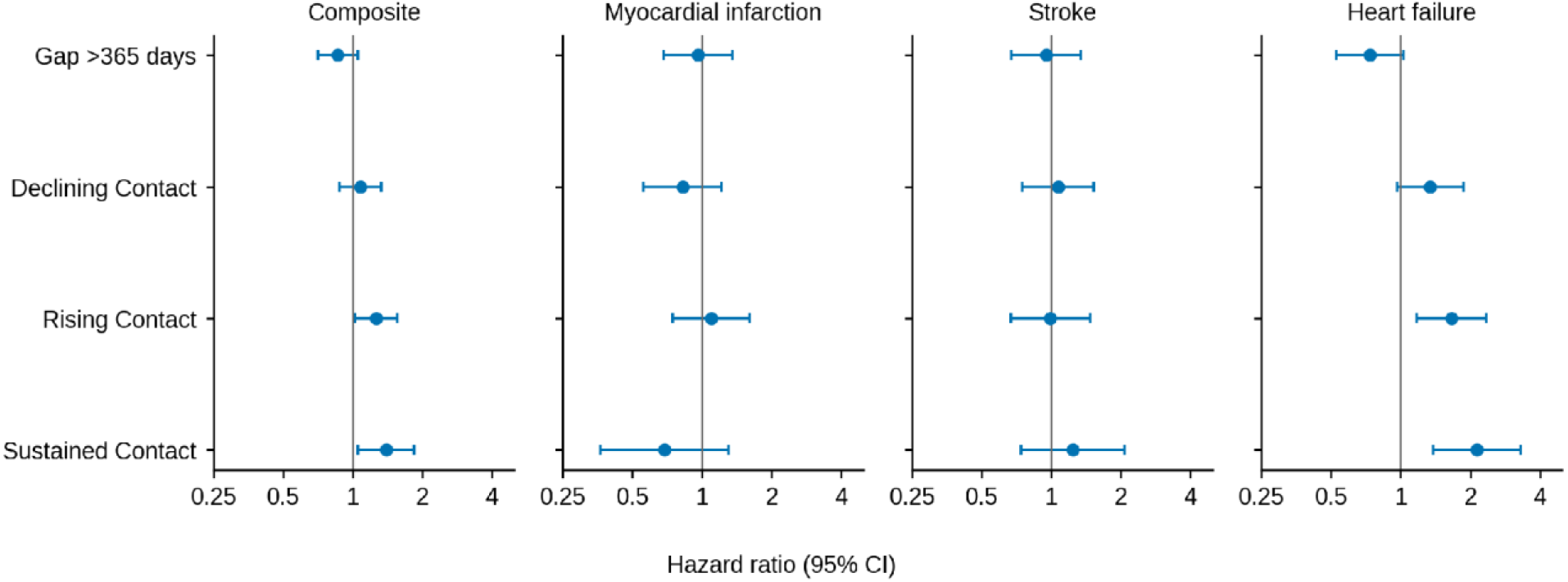
Adjusted associations of gap status and trajectory class with each outcome (exploratory). Cause-specific hazard ratios (95% CI) for a gap in recorded contact longer than 365 days (reference: no gap) and each trajectory class (reference: Minimal Contact), adjusted for the covariates in Table S7 and pooled over 30 imputations, for cardiovascular hospitalization (the composite) and for hospitalization with myocardial infarction, stroke, or heart failure as the principal diagnosis. Component outcomes are exploratory (12 comparisons: a gap longer than 365 days and three class contrasts, each for myocardial infarction, stroke, and heart failure hospitalization).

## REFERENCES

1. Ostchega, Y., et al., Hypertension prevalence among adults aged 18 and over: United States, 2017-2018. 2020: Hyattsville, MD.

2. Fuchs, F.D. and P.K. Whelton, High Blood Pressure and Cardiovascular Disease. Hypertension, 2020. 75(2): p. 285–292.

3. Muntner, P., et al., Trends in Blood Pressure Control Among US Adults With Hypertension, 1999-2000 to 2017-2018. JAMA, 2020. 324(12): p. 1190.

4. Ye, J., et al., Interventions and contextual factors to improve retention in care for patients with hypertension in primary care: Hermeneutic systematic review. Preventive Medicine, 2024: p. 107880.

5. Whelton, P.K., et al., 2017 ACC/AHA/AAPA/ABC/ACPM/AGS/APhA/ASH/ASPC/NMA/PCNA Guideline for the Prevention, Detection, Evaluation, and Management of High Blood Pressure in Adults: Executive Summary: A Report of the American College of Cardiology/American Heart Association Task F. Hypertension, 2018. 71(6): p. 1269–1324.

6. Lu, Y., et al., Delayed Hypertension Diagnosis and Its Association With Cardiovascular Treatment and Outcomes. JAMA Network Open, 2025. 8(7): p. e2520498.

7. Vickers, A.J. and E.B. Elkin, Decision Curve Analysis: A Novel Method for Evaluating Prediction Models. Medical Decision Making, 2006. 26(6): p. 565–574.

8. Hashish, M.A., S. Bronstein, and J. Ye, Returning value to communities from the All of Us Research Program through the lens of social determinants of health and ethical, legal, and social implications. International Journal for Equity in Health, 2026.

9. Obermeyer, Z., et al., Dissecting racial bias in an algorithm used to manage the health of populations. Science, 2019. 366(6464): p. 447–453.

10. Kleinberg, J., S. Mullainathan, and M. Raghavan. Inherent trade-offs in the fair determination of risk scores. in Proceedings of the 8th Innovations in Theoretical Computer Science Conference (ITCS). 2017.

11. Mugavero, M.J., et al., From Access to Engagement: Measuring Retention in Outpatient HIV Clinical Care. AIDS Patient Care and STDs, 2010. 24(10): p. 607–613.

12. Ye, J., et al., Characteristics and Patterns of Retention in Hypertension Care in Primary Care Settings From the Hypertension Treatment in Nigeria Program. JAMA Network Open, 2022. 5(9): p. e2230025–e2230025.

13. Nagin, D.S. and C.L. Odgers, Group-Based Trajectory Modeling in Clinical Research. Annual Review of Clinical Psychology, 2010. 6(1): p. 109–138.

14. Chen, H. and J. Ye, Machine Learning Prediction of Blood Pressure Control in Patients With Hypertension and Heart Failure Using Longitudinal Clinical Data. medRxiv, 2025: p. 2025.12. 25.25343007.

15. Choi, D., et al., Impact of continuity of care on cardiovascular disease risk among newly-diagnosed hypertension patients. Scientific Reports, 2020. 10(1).

16. Hernán, M.A., S. Hernández-Díaz, and J.M. Robins, A Structural Approach to Selection Bias. Epidemiology, 2004. 15(5): p. 615–625.

17. Hernán, M.A., et al., Specifying a target trial prevents immortal time bias and other self-inflicted injuries in observational analyses. Journal of Clinical Epidemiology, 2016. 79: p. 70–75.

18. Dafni, U., Landmark Analysis at the 25-Year Landmark Point. Circulation: Cardiovascular Quality and Outcomes, 2011. 4(3): p. 363–371.

19. Suissa, S., Immortal Time Bias in Pharmacoepidemiology. American Journal of Epidemiology, 2008. 167(4): p. 492–499.

20. Goldstein, B.A., et al., Controlling for Informed Presence Bias Due to the Number of Health Encounters in an Electronic Health Record. American Journal of Epidemiology, 2016. 184(11): p. 847–855.

21. Johnson, A.E.W., et al., MIMIC-IV, a freely accessible electronic health record dataset. Scientific Data, 2023. 10(1).

22. Johnson, A., et al., MIMIC-IV-ED, PhysioNet, Editor. 2023, 2.2.

23. Wang, X., Z. Ren, and J. Ye, Predicting survival time for critically ill patients with heart failure using conformalized survival analysis. AMIA Summits on Translational Science Proceedings, 2025. 2025: p. 576.

24. Chen, H. and J. Ye, Latent Class Trajectory Phenotypes of Longitudinal Visit and Follow-up Patterns Among Patients with Hypertension. medRxiv, 2026: p. 2026.07. 22.26358744.

25. Chen, T. and C. Guestrin. Xgboost: A scalable tree boosting system. in Proceedings of the 22nd acm sigkdd international conference on knowledge discovery and data mining. 2016.

26. Saito, T. and M. Rehmsmeier, The Precision-Recall Plot Is More Informative than the ROC Plot When Evaluating Binary Classifiers on Imbalanced Datasets. PLOS ONE, 2015. 10(3): p. e0118432.

27. Van Calster, B., et al., Calibration: the Achilles heel of predictive analytics. BMC Medicine, 2019. 17(1).

28. Lundberg, S.M. and S.-I. Lee. A unified approach to interpreting model predictions. in Advances in Neural Information Processing Systems 30 (NIPS 2017). 2017. Long Beach, CA, USA.

29. Collins, G.S., et al., Transparent reporting of a multivariable prediction model for individual prognosis or diagnosis (TRIPOD): the TRIPOD statement. BMJ, 2015. 350(jan07 4): p. g7594–g7594.

30. Hardt, M., E. Price, and N. Srebro. Equality of opportunity in supervised learning. in Advances in Neural Information Processing Systems 29 (NIPS 2016). 2016. Barcelona, Spain.

31. Proust-Lima, C., V. Philipps, and B. Liquet, Estimation of Extended Mixed Models Using Latent Classes and Latent Processes: The R Package lcmm. Journal of Statistical Software, 2017. 78(2).

32. Nagin, D.S., Group-Based Modeling of Development. 2005, Cambridge, MA: Harvard University Press.

33. Harrell, F.E., Jr., Regression Modeling Strategies: With Applications to Linear Models, Logistic and Ordinal Regression, and Survival Analysis. 2nd ed. 2015, Cham: Springer.

34. McCormick, N., et al., Validity of Heart Failure Diagnoses in Administrative Databases: A Systematic Review and Meta-Analysis. PLoS ONE, 2014. 9(8): p. e104519.

35. Quan, H., et al., Coding Algorithms for Defining Comorbidities in ICD-9-CM and ICD-10 Administrative Data. Medical Care, 2005. 43(11): p. 1130–1139.

36. Van Walraven, C., et al., A Modification of the Elixhauser Comorbidity Measures Into a Point System for Hospital Death Using Administrative Data. Medical Care, 2009. 47(6): p. 626–633.

37. White, I.R., P. Royston, and A.M. Wood, Multiple imputation using chained equations: Issues and guidance for practice. Statistics in Medicine, 2011. 30(4): p. 377–399.

38. White, I.R. and P. Royston, Imputing missing covariate values for the Cox model. Statistics in Medicine, 2009. 28(15): p. 1982–1998.

39. Rubin, D.B., Multiple Imputation for Nonresponse in Surveys. 1987, New York: Wiley.

40. Groenwold, R.H.H., et al., Missing covariate data in clinical research: when and when not to use the missing-indicator method for analysis. Canadian Medical Association Journal, 2012. 184(11): p. 1265–1269.

41. Carpenter, J.R. and M.G. Kenward, Multiple Imputation and its Application. 2013, Chichester: Wiley.

42. Gray, R.J., A class of K-sample tests for comparing the cumulative incidence of a competing risk. Annals of Statistics, 1988. 16: p. 1141–1154.

43. Fine, J.P. and R.J. Gray, A Proportional Hazards Model for the Subdistribution of a Competing Risk. Journal of the American Statistical Association, 1999. 94(446): p. 496–509.

44. Austin, P.C., D.S. Lee, and J.P. Fine, Introduction to the Analysis of Survival Data in the Presence of Competing Risks. Circulation, 2016. 133(6): p. 601–609.

45. Grambsch, P.M. and T.M. Therneau, Proportional hazards tests and diagnostics based on weighted residuals. Biometrika, 1994. 81(3): p. 515–526.

46. Cole, S.R. and M.A. Hernan, Constructing Inverse Probability Weights for Marginal Structural Models. American Journal of Epidemiology, 2008. 168(6): p. 656–664.

47. Austin, P.C. and E.A. Stuart, Moving towards best practice when using inverse probability of treatment weighting (IPTW) using the propensity score to estimate causal treatment effects in observational studies. Statistics in Medicine, 2015. 34(28): p. 3661–3679.

48. McCaffrey, D.F., et al., A tutorial on propensity score estimation for multiple treatments using generalized boosted models. Statistics in Medicine, 2013. 32(19): p. 3388–3414.

49. Uno, H., et al., On the C-statistics for evaluating overall adequacy of risk prediction procedures with censored survival data. Statistics in Medicine, 2011. 30(10): p. 1105–1117.

50. Schoenfeld, D.A., Sample-size formula for the proportional-hazards regression model. Biometrics, 1983. 39(2): p. 499–503.

51. Zhang, Z. and J. Ye, Predicting mortality in critically ill patients with hypertension using machine learning and deep learning models. Frontiers in Cardiovascular Medicine, 2025. 12: p. 1568907.

52. Carreras-García, D., et al., Patient No-Show Prediction: A Systematic Literature Review. Entropy, 2020. 22(6): p. 675.

53. Ye, J., et al., Community-Based Participatory Research and System Dynamics Modeling for Improving Retention in Hypertension Care. JAMA Network Open, 2024. 7(8): p. e2430213–e2430213.

54. Ye, J., et al., Implementation outcomes from the Hypertension Treatment in Nigeria program: results from a type 2 hybrid interrupted time series trial. Implementation Science, 2025.

55. Zhang, S., et al., Machine learning-based mortality prediction in critically ill patients with hypertension: comparative analysis, fairness, and interpretability. Frontiers in Artificial Intelligence, 2025. 8: p. 1686378.

56. Mugavero, M., et al., Missed Visits and Mortality among Patients Establishing Initial Outpatient HIV Treatment. Clinical Infectious Diseases, 2009. 48(2): p. 248–256.

57. Ye, J. and A. Song, Retention and loss to follow-up among patients with hypertension in primary care: a multi-practice cohort study. medRxiv, 2026: p. 2026.05. 10.26352856.

58. Ye, J., S. Bronstein, and M.A. Hashish, Learning health systems for cardiovascular health and health care. BMC Medicine, 2026.

59. Chen, H. and J. Ye, Demographic Patterns, Social Determinants of Health, and Climate Change Perceptions Among Patients with Cancer and Hypertension. medRxiv, 2026: p. 2026.01. 21.26344565.

